# Early-life nutritional environment is associated with late-life cognition in the Health and Retirement Study, a pellagra epidemic natural experiment

**DOI:** 10.64898/2026.06.11.26355481

**Authors:** Eva Vasiljevic, Lauren L. Schmitz, Corinne D. Engelman

**Affiliations:** Department of Population Health Sciences, University of Wisconsin School of Medicine and Public Health, 610 Walnut Dr., Madison, WI 53726, USA; Center for Demography of Health and Aging, University of Wisconsin-Madison, 1180 Observatory Drive Madison, WI 53706, USA; Robert M. La Follette School of Public Affairs, University of Wisconsin-Madison, 1225 Observatory Drive, Madison, WI 53706

## Abstract

Early-life exposures are important to several late-life health outcomes. We sought to study the effect of an *in utero* nutritional environment and its interaction with Alzheimer’s disease (AD) genetic risk on late-life cognitive function. We used a natural experiment created by the pellagra epidemic, a nutritional disease caused by a vitamin B_3_ deficiency, to evaluate the association between *in utero* pellagra epidemic exposure and late-life cognitive function in the Health and Retirement Study (N = 18,285). We also evaluated whether the *in utero* exposure could modify the AD polygenic score’s (PGS) effect on cognition. *In utero* pellagra epidemic exposure was significantly associated with cognition (β = −0.025). However, these effects were not isolated to the prenatal period as exposure during childhood periods also had an effect. The interaction between the *in utero* exposure and the AD PGS was significant, where the genetic effect on cognition was amplified with increasing (progressively worse) *in utero* exposure levels. These associations imply that the early-life nutritional environment affects late-life cognitive function and that these effects can modify genetic risk.

## 1.1 INTRODUCTION

Efforts to uncover the causes of Alzheimer’s disease (AD), the most common form of dementia, have focused on the pre-clinical stage of AD characterized by pathological and cognitive changes starting approximately 20 years before disease onset (Jack et al., 2013). Not much is known about the effects of events preceding midlife. Yet, early-life factors including *in utero* exposure to times of famine or influenza epidemics have links to cognitive function in late-life (de Rooij et al., 2010), and have been associated with multiple other late-life outcomes (Almond and Mazumder, 2005; de Rooij et al., 2021; Lumey et al., 2011). Studying what early-life factors lead to late-life cognitive changes is difficult because collecting data over such a long period of time is challenging. In the absence of data over the entire life course, natural experiments are one tool to evaluate relationships between early-life experiences and late-life health outcomes. Our study takes advantage of a natural experiment created by the pellagra epidemic (a nutritional disease caused by a vitamin B_3_ deficiency) in the early 20th century United States (US) to study the potential effects of the early-life nutritional environment on late-life cognitive function and whether this early-life exposure modifies genetic risk of AD.

Nutrition is one modifiable factor of interest in dementia research because it affects neural development (Cusick and Georgieff, 2016; Goyal et al., 2018) and because associations between dietary patterns and cognitive function and AD risk have been identified (Morris et al., 2015; Samadi et al., 2019). Vitamin B_3_, in particular, has connections to neural health and cognitive function. Severe deficiencies lead to the dementia-related disease pellagra (Goyal et al., 2018); treatment with vitamin B_3_ promotes remyelination of cultured human and mice microglia (Rawji et al., 2020); and higher dietary intakes of vitamin B_3_ are associated with slower rate of cognitive decline and lower dementia risk (Morris et al., 2004). In addition, interactions between genetic risk of AD and diet have been of interest (Martínez-Lapiscina et al., 2014; Samuelsson et al., 2022). Understanding whether a modifiable factor such as diet can mitigate genetic risk of dementia and not only overall risk is an important tool in uncovering how to reduce dementia burden. These and most studies about the relationship between diet and dementia or cognition examine mid- or late-life diet and do not address early-life diet given the difficulty of measuring early-life exposures for older cohorts. In addition, observational studies have challenges including the use of self-report for food intake, which is affected by poor recall, and disentangling whether diet affects cognition or whether they share a common cause such as access to resources (Satija et al., 2015). This leaves a need to not only evaluate the effects of early-life nutrition on late-life cognitive function but to do so in a way that is less prone to spurious associations or poor recall. We address that need by using a natural experiment created by the pellagra epidemic in the early 20^th^ century US.

Pellagra is a disease caused by a deficiency in vitamin B_3_ (also called niacin), the primary dietary sources of which are meats, legumes, and whole grains (Barrett-Connor, 1967; Office of Dietary Supplements, National Institutes of Health, 2021). It is a reversible condition characterized by the four “Ds”: diarrhea, dermatitis, dementia, and death if untreated (Lanska, 1996). Changes in the grain milling process and treatment of corn in the late 19th century affected the nutritional content of cereal-derived foods such as bread and flour, effectively removing certain B vitamins (Brenton, 2000). Consequently, the US experienced a pellagra epidemic, with estimates that for every death there were 35 non-fatal cases between 1915 and 1925 (Bollet, 1992). Pellagra was more prevalent among women and Black populations, and it was in the top ten leading causes of death in southern states (Brenton, 2000; DeKleine, 1942; Marks, 2003). After uncovering treatments that alleviated pellagra and identified its cause (Barrett-Connor, 1967), there were voluntary and mandatory responses to the epidemic and general malnutrition. Individual states passed laws requiring bakers and millers to fortify bread and flour (Park et al., 2001) with vitamins B_1_, B_2_, B_3_, iron, and, optionally, calcium (Wilder, 1956). By 1955, the pellagra epidemic was over thanks to these and other nutritional improvements (Park et al., 2000). The staggered response to the epidemic created variation across the nation in who had access to B vitamin enriched foods, which consequently led to variation in pellagra death rates across states and time. This variation occurred in the early-life years of participants in the Health and Retirement Study (HRS) and created an opportunity to measure differences in the early-life nutritional environment.

In this study, we leverage time and place-of-birth data from the HRS to evaluate the association between state of birth and gestation year pellagra mortality rates, which reflect the *in utero* nutritional environment, and late-life cognition. We also evaluate whether *in utero* exposure to the pellagra epidemic modifies the effect of AD genetic risk on late-life cognition. Additionally, we evaluate time periods during childhood to see if other early-life periods influence late-life cognition. This will help identify important time points in the life course that affect cognition and will provide evidence on the role of the early-life nutritional environment that are less prone to typical methodological challenges of studying a complex behavioral exposure.

### 1.2 METHODS

#### 1.2.1 Sample and data

##### HRS description

HRS is a nationally representative longitudinal study of pre-retirement and retirement aged US adults that began in 1992 (Heeringa and Connor, 1995). This project used public, sensitive, and restricted HRS data from the years 1992–2020, with more specific information provided in **Supplemental Methods**.

##### Analytic dataset

From a starting sample of 42,405 individuals and 280,343 observations of completed HRS core interviews, we excluded observations with missing values for the outcome (27-point cognition), exposure (*in utero* pellagra epidemic exposure), and covariates. Exposure missingness is related to both individuals’ availability of month and year of birth data and year and state pellagra mortality statistic availability. After initial exclusions, 19,283 individuals and 140,904 observations remained. We also removed observations (not participants) post self-reported stroke and observations before the age of 50, since cognitive change due to dementia before this age is rare (Barnes et al., 2015). After all exclusions, 18,285 individuals and 128,927 observations remained. The analytic dataset included up to 15 observations per person with a median of 8 observations. With an average time of 2.1 years between observations, this reflects a median time of 13 and maximum of >18 years in the study. Additional analyses were conducted with genetic variables, with an analytic sample of 7,625 individuals and 71,250 observations after removing individuals with missing values for the AD PGS, of non-European genetic ancestry, and with apolipoprotein E (*APOE*) variant imputation quality scores lower than 0.8 (Casaburi et al., 2025).

#### 1.2.2 Measures

##### Exposure: Early-life pellagra epidemic exposure

The pellagra mortality per 100,000 people (100K) for an individual’s state of birth and their gestation year was the exposure of interest, referred to as the “*in utero* pellagra epidemic exposure.” For participants whose gestation spanned more than one calendar year, a weighted average of the pellagra mortality rates for the two years was used, weighted by the number of months gestating in each year. Higher rates were considered worse environmental conditions. By the nature of the data assembled in this study, this measure was only available for people born in the US, and in states and years that reported pellagra deaths (1915–1950, **Supplemental Figure 1**). In addition, this was not a direct measure of a person’s dietary exposure to vitamin B_3_ or to other B vitamins that foods were fortified with in response to the epidemic. This is also not a measure of the rate of deaths *in utero*, rather it is the environmental conditions in states and years with differing rates of pellagra mortality during the epidemic. State of birth was obtained from HRS restricted geographic data. Additional measures include exposure to the pellagra epidemic during the childhood periods of 0–2, 3–6, and 7–10 years, which is the average of the pellagra mortality rates in the state of birth for each participant over the calendar years a participant was a given age. More information on the public mortality data sources and the derivation of the exposure measures are detailed in **Supplemental Table 1** and the **Supplemental Methods**.

##### Outcome: Global cognition

The main outcome in the study was global cognition on a 27-point scale obtained from the researcher-contributed HRS dataset “Langa-Weir Classification of Cognitive Function,” based on a modified version of the Telephone Interview for Cognitive Status (TICS) (Ofstedal et al., 2005). The 35-point TICS global cognition measure was used in sensitivity analyses and was only administered to individuals 65 years of age or older, which is why it is not used as the primary analysis outcome. Higher values indicate better cognition for both measures. Components of each cognitive measure are listed in the **Supplemental Methods**.

##### Genetic measures

The main genetic exposure in the analyses was the PGS for AD (including *APOE* variants). A PGS is a summary measure of genetic risk from multiple variants across the genome. Higher values of the AD PGS indicate higher risk of AD. The score was generated using 39 variants (**Supplemental Table 2**) as reported in de Rojas et al., 2021 and 2 *APOE* variants (rs7412 and rs429358), using weights from genome-wide association studies (GWAS) conducted in individuals of European genetic ancestry.

Because the weights were developed for individuals of European genetic ancestry, PGSs were only generated for individuals with that genetic ancestry. More information about the PGS generation Supplemental Methodsand additional genetic measures (including an *APOE* score, an alternative HRS-provided AD PGSs with and without *APOE*, and an HRS-provided PGS for cognition) are described in the **Supplemental Methods**. Unlike the AD PGS, higher values of the cognition PGS indicated better cognition.

##### Covariates

Additional covariates included: participant’s age at the last interview date (centered at 65), gender, race, participant’s education, mother’s education, childhood family financial situation, birth year cohort, birth Census division, interview language, and cognitive testing practice effects. HRS categorizes race as White, Black and other (aggregated to protect participant identity in smaller groups). Genetic analyses also adjusted for population stratification (e.g., confounding due to genetic ancestry) by including 10 genetic principal components, provided by HRS. Sensitivity analyses also included binary indicators of ever having had a stroke, hypertension, heart disease or problems, diabetes, and smoking history. More details about the covariates are included in the **Supplemental Methods**.

#### 1.2.3 Statistical analysis

##### In utero pellagra epidemic exposure association with cognition

We used mixed effects regression to evaluate the association of the *in utero* pellagra epidemic exposure with the main cognitive outcome (27-point global cognition). We included a random intercept at the individual level to account for repeated measures correlation.

The first model was the crude association of the *in utero* exposure with global cognition. The second model added birth year cohort and birth location. The third model added age, gender, race, mother’s education, participant’s education, practice effects, interview language, and childhood family financial situation; age was included as a cubic polynomial after confirming it improved model fit over linear age using the Akaike Information Criterion. This final model was used as the basis in subsequent analyses unless otherwise specified. Additional regression modeling information is included in the **Supplemental Methods**.

##### Stratification by race and gender

The analysis was stratified by race and gender to examine how the effects varied by these two factors that are both related to differential underlying experience of the pellagra epidemic and cognitive testing performance. For these analyses, interview language was not adjusted for because not all strata had a mixture of English and Spanish interviews. The full sample model without adjusting for interview language was included as a comparison.

##### Interaction between PGS and in utero pellagra epidemic exposure

To test whether *in utero* pellagra epidemic exposure can modify a person’s genetic risk of AD, we included an interaction term between the AD PGS (that includes *APOE*) with the *in utero* exposure in the fully-adjusted model, excluding race and interview language, which did not have any variation in this subsample. Because variants in *APOE* are among the strongest genetic risk predictors for AD and can account for much of the effect of a genetic association, sensitivity analyses were conducted excluding *APOE* variants from the PGS and testing *APOE* risk alone. We also tested the HRS-provided AD PGSs with and without *APOE* variants generated for individuals of European genetic ancestry. In addition, we conducted analyses using the PGS for cognition. In another sensitivity analysis, we added an interaction between each principal component and the *in utero* pellagra epidemic exposure.

##### Exposures to the pellagra epidemic during childhood

Childhood is another period of development that can be sensitive to environmental influences that carry into adulthood. To test whether exposure to the pellagra epidemic during childhood had effects on late-life cognitive function, we individually evaluated the effects of living through to the pellagra epidemic on global cognition of three additional age periods: 0–2, 3–6 and 7–10 years. We also tested the three age periods in a single regression model with *in utero* exposure to the pellagra epidemic to see if any one age period had a strong effect over others on cognition.

##### Additional sensitivity analyses

We performed sensitivity analyses to evaluate whether the association of *in utero* pellagra epidemic exposure varies under different conditions. First, we repeated the fully-adjusted analysis and the genetic interaction analysis using a different outcome, the 35-point global cognition score.

Second, we determined whether the association between *in utero* pellagra epidemic exposure and cognition and the interaction between *in utero* pellagra epidemic exposure and the AD PGS was consistent if the analysis was conducted in a subsample of individuals born in the US South Census region, where higher mortality from the pandemic was observed (**Supplemental Figure 2**). Conducting an analysis within this stratum accounts for potential interaction effects with being born in the South that are not captured in the main model. Analyses in the other three regions were not conducted because low pellagra epidemic severity risked violating the positivity assumption.

Third, we evaluated whether alternative specifications of the birth location and time would affect the main exposure’s estimates. In the main analyses, the models were adjusted for Census division of birth (10 categories) and birth year in five-year intervals (8 categories), which are aggregate categories of the state of birth and birth year. Those relatively large categories may mask important variation in state of birth and birth year differences. To account for that, these terms were specified in two alternative ways. One method used fixed effects for individual states of birth instead of Census division categories and individual birth years instead of five-year categories. The other method included random intercepts for state of birth and for birth year in the model.

Finally, we reanalyzed the association between *in utero* pellagra epidemic exposure and cognition in additional ways. The first removed participant’s education from models. Education was conceptually assumed to be a confounder but could have a mediating effect, so we present models without education. Likewise, in a separate analysis, we adjusted for history of five health events or conditions (history of stroke, hypertension, heart disease or problems, diabetes, and smoking) that may affect cognition.

#### 1.2.4 Software

HRS data were accessed and analyzed on the University of Michigan’s Michigan Center on the Demography and Aging virtual data enclave. The statistical software R (v 4.4.1) was used for all analyses (R Core Team, 2020). Additional R packages were used for regression analysis (Bates et al., 2015; Kuznetsova et al., 2017), model parameter extraction (Lüdecke et al., 2021, 2020), estimation of model predicted values for interactions (Lenth, 2020), and data management and visualization (Wickham et al., 2019). PLINK (v 1.9) was used for processing individual-level genetic data (Chang et al., 2015; Purcell and Chang, n.d.).

#### 1.2.5 Ethics

HRS obtained permission to conduct the study through the University of Michigan Institutional Review Board (IRB). HRS participants provide informed consent before each interview. Secondary data analysis was approved by the University of Wisconsin-Madison IRB.

### 1.3 RESULTS

#### 1.3.1 Sample characteristics

The mean age at first cognitive assessment was 62 years for the full analytic sample and 61 years for the genetic analyses sample (**Table 1**). The sample was primarily composed of participants who identified as White in the full sample and only composed of participants who identified as White in the genetic analysis sample. Genetic analyses used PGSs with weights from GWAS conducted in individuals with European genetic ancestry. These PGSs were only available for HRS participants with European genetic ancestry, which corresponded to participants who also identified as White in this sample. Differences in sample composition when stratified by the exposure reflected differences in where and when the pellagra epidemic was most severe. The South Census region and birth years between approximately 1921 and 1941 had the highest number of individuals in the higher exposure stratum. In addition, those with higher exposure to the pellagra epidemic were on average older at the time of interview than those with lower exposure to the pellagra epidemic.

**Table 1:** Participant characteristics in the full analytic sample, in the genetic analysis sample, and stratified by the exposure of interest within each.

| Characteristic | Full sample |  |  | Genetic analyses |  |  |
| --- | --- | --- | --- | --- | --- | --- |
|  | Total<br>(N = 18285) | By exposure strata, *in utero*<br>pellagra epidemic exposure<br>(mortality rate per 100k) |  | Total<br>(N = 7625) | By exposure strata, *in utero*<br>pellagra epidemic exposure<br>(mortality rate per 100k) |  |
|  |  | <= 10<br>(N = 16149) | > 10<br>(N = 2136) |  | <= 10<br>(N = 7083) | > 10<br>(N = 542) |
| **State and year of gestation pellagra mortality rate per 100k (mean, SD)** | 3 (6) | 1 (2) | 17 (8) | 2 (5) | 0.89 (1.90) | 16 (7) |
| **Global cognition, 27-point (mean, SD)<sup>a</sup>** | 16 (4) | 16 (4) | 14 (5) | 17 (4) | 17 (4) | 17 (4) |
| **Global cognition, 35-point (mean, SD)<sup>a</sup>** | 24 (5) | 24 (5) | 21 (6) | 25 (4) | 25 (4) | 24 (4) |
| **Age, years (mean, SD)** | 62 (8) | 62 (8) | 66 (6) | 61 (8) | 60 (8) | 65 (6) |
| **Female (n, %)** | 10234 (56%) | 8991 (56%) | 1243 (58%) | 4332 (57%) | 4010 (57%) | 322 (59%) |
| **Self-reported race (n, %)<sup>b</sup>** |  |  |  |  |  |  |
| White or caucasian | 15020 (82%) | 13828 (86%) | 1192 (56%) | 7625 (100%) | 7083 (100%) | 542 (100%) |
| Black or AA | 2807 (15%) | 1914 (12%) | 893 (42%) | 0 (0%) | 0 (0%) | 0 (0%) |
| Other | 458 (3%) | 407 (3%) | 51 (2%) | 0 (0%) | 0 (0%) | 0 (0%) |
| **Education (n, %)** |  |  |  |  |  |  |
| Less than HS | 3871 (21%) | 3019 (19%) | 852 (40%) | 986 (13%) | 888 (13%) | 98 (18%) |
| GED | 857 (5%) | 737 (5%) | 120 (6%) | 339 (4%) | 305 (4%) | 34 (6%) |
| HS graduate | 6026 (33%) | 5445 (34%) | 581 (27%) | 2656 (35%) | 2461 (35%) | 195 (36%) |
| Some college | 3975 (22%) | 3686 (23%) | 289 (14%) | 1767 (23%) | 1670 (24%) | 97 (18%) |
| College and above | 3556 (19%) | 3262 (20%) | 294 (14%) | 1877 (25%) | 1759 (25%) | 118 (22%) |
| **Mother's education (n, %)** |  |  |  |  |  |  |
| No Schooling | 281 (2%) | 237 (1%) | 44 (2%) | 29 (0%) | 27 (0%) | suppressed<sup>c</sup> |
| Less than 8 yrs | 3235 (18%) | 2594 (16%) | 641 (30%) | 966 (13%) | 839 (12%) | suppressed<sup>c</sup> |
| Completed 8 yrs | 6284 (34%) | 5570 (34%) | 714 (33%) | 2752 (36%) | 2535 (36%) | suppressed<sup>c</sup> |
| HS graduate | 4555 (25%) | 4308 (27%) | 247 (12%) | 2320 (30%) | 2228 (31%) | suppressed<sup>c</sup> |
| Above HS | 1832 (10%) | 1708 (11%) | 124 (6%) | 1025 (13%) | 973 (14%) | suppressed<sup>c</sup> |
| Missing | 2098 (11%) | 1732 (11%) | 366 (17%) | 533 (7%) | 481 (7%) | suppressed<sup>c</sup> |
| **Childhood family financial situation (n, %)** |  |  |  |  |  |  |
| Poor | 5446 (30%) | 4589 (28%) | 857 (40%) | 2099 (28%) | 1893 (27%) | 206 (38%) |
| Average | 10861 (59%) | 9766 (60%) | 1095 (51%) | 4911 (64%) | 4592 (65%) | 319 (59%) |
| Well off | 1069 (6%) | 1008 (6%) | suppressed<sup>c</sup> | suppressed<sup>c</sup> | suppressed<sup>c</sup> | suppressed<sup>c</sup> |
| Varied | 179 (1%) | 169 (1%) | suppressed<sup>c</sup> | suppressed<sup>c</sup> | suppressed<sup>c</sup> | suppressed<sup>c</sup> |
| Missing | 730 (4%) | 617 (4%) | suppressed<sup>c</sup> | suppressed<sup>c</sup> | suppressed<sup>c</sup> | suppressed<sup>c</sup> |
| **Stroke, ever (n, %)** | 184 (1%) | 158 (1%) | 26 (1%) | 91 (1%) | 79 (1%) | 12 (2%) |
| **Hypertension, ever (n, %)** | 12784 (70%) | 11162 (69%) | 1622 (76%) | 5485 (72%) | 5059 (71%) | 426 (79%) |
| **Heart disease or problems, ever (n, %)** | 7015 (38%) | 6111 (38%) | 904 (42%) | 3284 (43%) | 3014 (43%) | 270 (50%) |
| **Diabetes, ever (n, %)** | 5224 (29%) | 4555 (28%) | 669 (31%) | 2068 (27%) | 1924 (27%) | 144 (27%) |
| **Interview language (n, %)** |  |  |  |  |  |  |
| English | 18195 (100%) | 16073 (100%) | 2122 (99%) | 7625 (100%) | 7083 (100%) | 542 (100%) |
| Spanish | 90 (0%) | 76 (0%) | 14 (1%) | 0 (0%) | 0 (0%) | 0 (0%) |
| **Birth year cohort (n, %)<sup>d</sup>** |  |  |  |  |  |  |
| 1911–1920 | 1296 (7%) | 1108 (7%) | 188 (9%) | 290 (4%) | 271 (4%) | 19 (4%) |
| 1921–1930 | 4057 (22%) | 3356 (21%) | 701 (33%) | 1598 (21%) | 1405 (20%) | 193 (36%) |

| (Continued) | Full sample |  |  | Genetic analyses |  |  |
| --- | --- | --- | --- | --- | --- | --- |
|  | By exposure strata, *in utero* pellagra epidemic exposure (mortality rate per 100k) |  |  | By exposure strata, *in utero* pellagra epidemic exposure (mortality rate per 100k) |  |  |
|  | Total<br>(N = 18285) | <= 10<br>(N = 16149) | > 10<br>(N = 2136) | Total<br>(N = 7625) | <= 10<br>(N = 7083) | > 10<br>(N = 542) |
| Characteristic |  |  |  |  |  |  |
| 1931–1940 | 6975 (38%) | 5728 (35%) | 1247 (58%) | 3022 (40%) | 2692 (38%) | 330 (61%) |
| 1941–1950 | 5770 (32%) | 5770 (36%) | 0 (0%) | 2639 (35%) | 2639 (37%) | 0 (0%) |
| 1951–1960 | 187 (1%) | 187 (1%) | 0 (0%) | 76 (1%) | 76 (1%) | 0 (0%) |
| **Birth Census region (n, %)<sup>e</sup>** |  |  |  |  |  |  |
| Midwest | 5638 (31%) | 5638 (35%) | suppressed<sup>c</sup> | 2902 (38%) | 2902 (41%) | 0 (0%) |
| Northeast | 4175 (23%) | 4175 (26%) | suppressed<sup>c</sup> | 1901 (25%) | 1901 (27%) | 0 (0%) |
| South | 6721 (37%) | 4588 (28%) | 2133 (100%) | 2079 (27%) | 1537 (22%) | 542 (100%) |
| West | 1751 (10%) | 1748 (11%) | suppressed<sup>c</sup> | 743 (10%) | 743 (10%) | 0 (0%) |

Note: Mean values for longitudinal variables (cognition measures and age) are from the baseline observation in the analytic dataset. For health history factors (stroke, hypertension, heart disease or problems, diabetes, and smoking), the presented summary values are based on ever having reported the factor across all visits.
<sup>b</sup> Genetic analyses used polygenic scores (PGSs) with weights from genome-wide association studies conducted in individuals with European genetic ancestry. These PGSs were only available for HRS participants with European genetic ancestry, which happened to correspond to participants who identified as White.
<sup>c</sup> Values are suppressed to mask one or more cells that have a count below 11 to preserve participant identities.
<sup>d</sup> Regression models are adjusted for 5-year birth cohort intervals instead of the 10-year intervals show here.
<sup>e</sup> Regression models are adjusted for birth Census division, which are smaller geographic groupings than the Census region.

#### 1.3.2 *In utero* pellagra epidemic exposure effects on global cognition

##### Main effects of pellagra epidemic exposure

*In utero* pellagra epidemic exposure was significantly associated with 27-point global cognition across all models in the full sample (**Table 2**, **Supplemental Table 3**). The effect of the main exposure attenuated after the addition of covariates from −0.148 (95% confidence interval [CI]: −0.156 to −0.139) in the unadjusted model to −0.025 (95% CI: −0.035 to −0.015) in the fully adjusted model, which meant that for every pellagra death per 100K increase in epidemic severity, mean global cognition was lower by 0.025 points. This effect size equated to an approximate one-point difference in global cognition between someone with no exposure (0 deaths per 100K) to someone with a high level of exposure (50 deaths per 100K), holding all other covariates constant.

**Table 2:** Association between *in utero* pellagra epidemic exposure and 27-point global cognition using mixed effects regression models.

| Covariate | Outcome: 27-point global cognition |  |  |
| --- | --- | --- | --- |
|  | Model 1<sup>a</sup> | Model 2<sup>b</sup> | Model 3<sup>c</sup> |
| Intercept | 15.618<br>(15.560 to 15.676) | 13.006<br>(12.761 to 13.251) | 11.676<br>(11.239 to 12.112) |
| *In utero* pellagra epidemic exposure | -0.148***<br>(-0.156 to -0.139) | -0.059***<br>(-0.070 to -0.047) | -0.025***<br>(-0.035 to -0.015) |
| Marginal R<sup>2</sup> / Conditional R<sup>2</sup> | 0.03 / 0.56 | 0.09 / 0.56 | 0.32 / 0.64 |
| AIC | 687648 | 685895 | 661621 |
| N people | 18285 | 18285 | 18285 |
| N observations | 128927 | 128927 | 128927 |

\* $p \leq 0.05$ \*\* $p \leq 0.01$ \*\*\* $p \leq 0.001$
Abbreviations: AIC: Akaike information criterion; N: number.
Notes: Values presented are coefficient estimates with 95% confidence intervals. All models include a random intercept at the person level. *In utero* pellagra epidemic exposure refers to the pellagra mortality rate during a participant's gestation year in their state of birth.
<sup>a</sup> Model 1 is unadjusted.
<sup>b</sup> Model 2 includes all terms from model 1 and adjusts for person's birth Census division and birth year cohort in 5-year intervals.
<sup>c</sup> Model 3 includes all terms from model 2 and adjusts for age (cubic polynomial centered at 65), gender, race, mother's education, participant's education, practice effects, interview language, and childhood family financial situation.

##### Stratified by gender and race

The association between *in utero* pellagra epidemic exposure and global cognition was no longer significant in all sub-samples except for Black females and females of other racial backgrounds (**Figure 1**, **Supplemental Table 4**). However, the magnitude and direction of the effect were consistent with the full sample effect (negative) for all strata except for Black males (β = 0.004, 95% CI: −0.027 to 0.035) and males of other racial backgrounds (β = 0.061, 95% CI: −0.072 to 0.194). *In utero* pellagra epidemic exposure associations with global cognition were approximately the same in White male and White female strata (β White male = −0.018, 95% CI: −0.036 to 0.001 and β White female = −0.015, 95% CI: −0.033 to 0.002). The exposure had the second-strongest negative effect in the Black female stratum (β = −0.031, 95% CI: −0.059 to −0.004) and the strongest negative effect in the other racial background female stratum (β = −0.145, 95% CI: −0.242 to −0.049), which was about a 5- to 10-fold greater magnitude in effect than in Black and White female strata.

**Figure 1:**
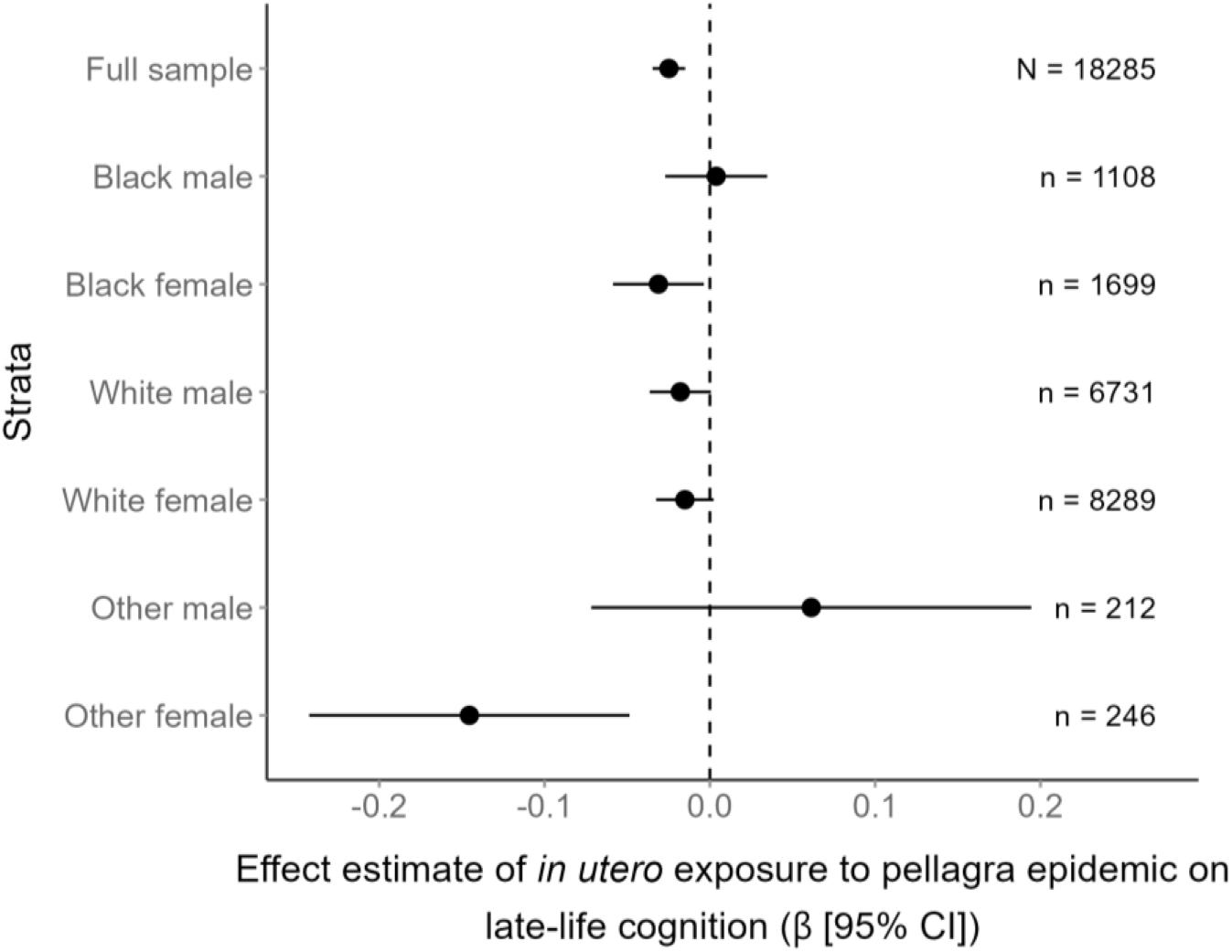
*In utero* pellagra epidemic exposure coefficient estimates (points) and 95% confidence intervals (lines) from analyses stratified by both racial background and gender. The outcome is 27-point cognition, and *in utero* pellagra epidemic exposure is measured by the pellagra mortality rates in a participant’s state of birth during their gestation year. Models were adjusted for age (cubic polynomial centered at 65), mother’s education, participant’s education, practice effects, childhood family financial situation, person’s birth Census division, and birth year cohort in 5-year intervals. Interview language was not included as a covariate (including in the full sample estimate presented here) because not all strata had mixtures of English and Spanish interviews. Other is an HRS-provided category that refers to a racial background that is not captured by either the Black or White racial background groups.

#### 1.3.3 Interaction between *in utero* pellagra epidemic exposure and genetics

The association between *in utero* pellagra epidemic exposure and global cognition was no longer significant in the sample of individuals with genetic data, with a magnitude of effect of −0.010 (95% CI: −0.027 to 0.006, model 1, **Table 3**). The interaction between *in utero* pellagra epidemic exposure and the AD PGS with *APOE* was significant (model 3, **Table 3** β = −1.915, 95% CI: −3.226 to −0.603). This equated to greater differences in the effect of genetics on cognition under more severe pellagra epidemic conditions (higher *in utero* pellagra epidemic exposure) compared to less severe conditions (lower exposure) (**Figure 2**). Stated another way, the negative environment amplified the genetic effect.

**Figure 2:**
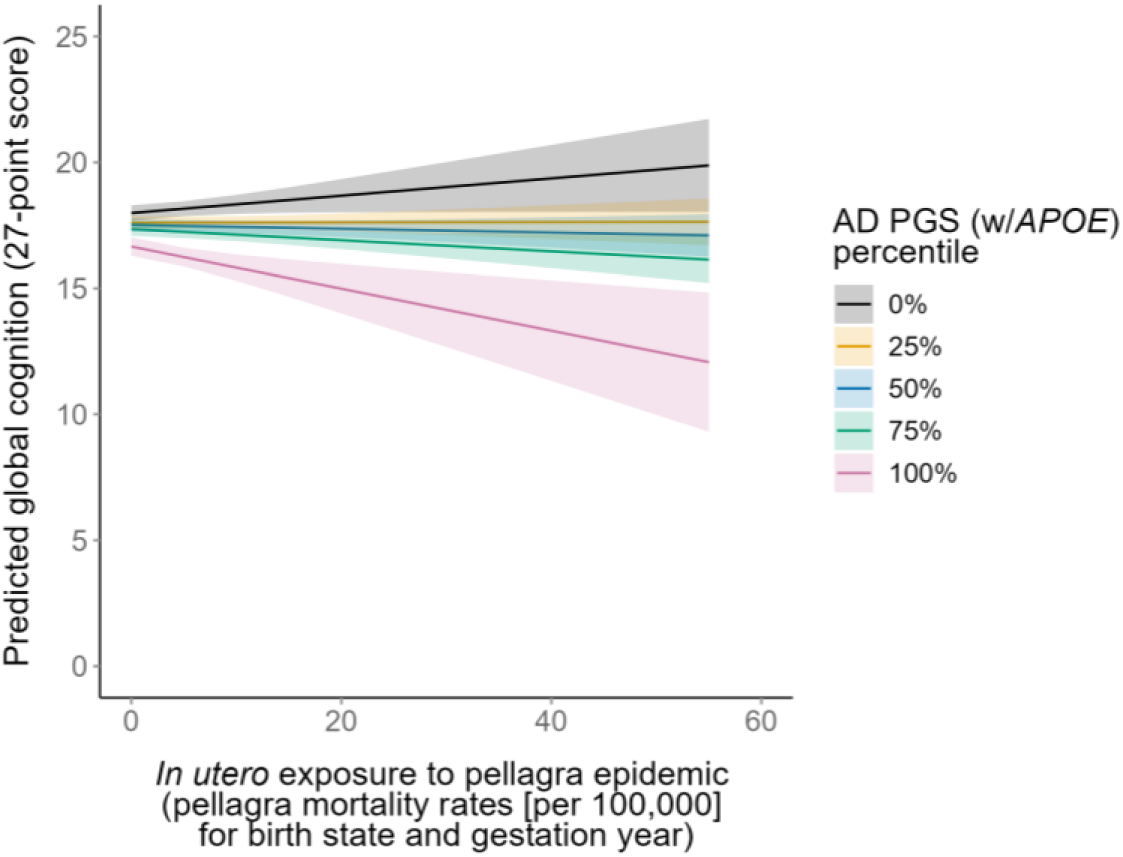
Interaction plot of AD PGS with *APOE* percentiles across levels of *in utero* pellagra epidemic exposure (x-axis) for 27-point global cognition (y-axis). Bands represent 95% confidence intervals. Estimates are plotted with fixed values for gender (female), mother’s education (completed 8 years), participant’s education (high school graduate), age (65 years), practice effects (2 counts), childhood family financial situation (“average”), birth Census division (South Atlantic), birth year cohort (1930–1934), and the sample mean of each principal component. Higher values of the PGS indicate higher risk of AD. Lower values of global cognition indicate poorer cognition. Higher values of the *in utero* pellagra epidemic exposure (state of birth and gestation year pellagra mortality rates) are representative of a worse environment.

**Table 3:** Effect estimates of the interaction between *in utero* pellagra epidemic exposure and the AD PGS on 27-point global cognition.

| Covariate | Outcome: 27-point global cognition |  |  |
| --- | --- | --- | --- |
|  | Model 1<sup>a</sup> | Model 2<sup>b</sup> | Model 3<sup>c</sup> |
| Intercept | 13.778<br>(12.743 to 14.813) | 13.799<br>(12.764 to 14.833) | 13.794<br>(12.760 to 14.828) |
| *In utero* pellagra epidemic exposure | -0.010<br>(-0.027 to 0.006) | -0.011<br>(-0.028 to 0.005) | -0.008<br>(-0.025 to 0.009) |
| AD PGS with *APOE* |  | -25.547***<br>(-31.681 to -19.413) | -21.689***<br>(-28.365 to -15.012) |
| PGS x *in utero* |  |  | -1.915**<br>(-3.226 to -0.603) |
| Marginal R<sup>2</sup> / Conditional R<sup>2</sup> | 0.27 / 0.58 | 0.27 / 0.58 | 0.27 / 0.58 |
| AIC | 357683 | 357587 | 357579 |
| N people | 7625 | 7625 | 7625 |
| N observations | 71250 | 71250 | 71250 |

\* p ≤ 0.05 \*\* p ≤ 0.01 \*\*\* p ≤ 0.001
Abbreviations: AD: Alzheimer's disease; AIC: Akaike information criterion; *APOE*: apolipoprotein E; N: number; PGS: polygenic score.
Notes: All models included a random intercept at the person level. Values presented are coefficient estimates with 95% confidence intervals. Models were adjusted for gender, age (cubic polynomial centered at 65), mother's education, participant's education, practice effects, childhood family financial situation, person's birth Census division, birth year cohort in 5-year intervals, and the first 10 principal components. *In utero* pellagra epidemic exposure refers to the pellagra mortality rate during a participant's gestation year in their state of birth.
<sup>a</sup> Model 1 is the main association of the *in utero* pellagra epidemic exposure with global cognition in the PGS analysis subsample.
<sup>b</sup> Model 2 models the addition of the AD PGS (with *APOE*).
<sup>c</sup> Model 3 tests the interaction between the AD PGS and the *in utero* pellagra epidemic exposure.

The interaction term was no longer significant in sensitivity analyses using the AD PGS that did not contain *APOE* (β = −1.288, 95% CI: −3.974 to 1.397, **Supplemental Table 5**) but was significant when *APOE* by itself was interacted with the exposure of interest (β = −0.028, 95% CI: −0.047 to −0.009, **Supplemental Table 5**). Adding an interaction between the *in utero* pellagra epidemic exposure and each principal component did not substantively affect the main interaction effect (β = −1.909, 95% CI: −3.235 to −0.583, **Supplemental Table 5**).

#### 1.3.4 Exposure to the pellagra epidemic during childhood

When tested individually, exposure to the pellagra epidemic during all three age periods was significantly associated with global cognition (**Table 4**). The effect sizes were similar to the *in utero* pellagra epidemic exposure (–0.025, model 1, **Table 2**) but slightly higher with age 0–2 having an effect size of −0.028 (model 2, 95% CI: −0.040 to −0.015); age 3–6 an effect of −0.036 (model 3, 95% CI: −0.050 to −0.022); and age 7–10 an effect of −0.038 (model 4, 95% CI: −0.054 to −0.022). However, when modeled together, none of the periods were significantly associated with global cognition (model 5, **Table 4**), and no single age period stood out as an influential period on cognitive function. Each pellagra epidemic exposure measure was highly correlated with one another, having Pearson correlations between 0.74–0.95. The high correlation reduced but did not eliminate the ability to isolate if exposure during one particular age period was the main influence on cognitive function in later life.

**Table 4:** Association of childhood pellagra epidemic exposure and 27-point global cognition.

| Covariate | Outcome: 27-point global cognition |  |  |  |  |
| --- | --- | --- | --- | --- | --- |
|  | Model 1 | Model 2 | Model 3 | Model 4 | Model 5 |
| Intercept | 11.337<br>(10.834 to 11.840) | 11.324<br>(10.822 to 11.827) | 11.326<br>(10.823 to 11.829) | 11.377<br>(10.873 to 11.880) | 11.919<br>(11.406 to 12.433) |
| *In utero* pellagra epidemic exposure | -0.025***<br>(-0.036 to -0.014) |  |  |  | -0.024<br>(-0.051 to 0.004) |
| Age 0–2 pellagra epidemic exposure |  | -0.028***<br>(-0.040 to -0.015) |  |  | 0.023<br>(-0.016 to 0.062) |
| Age 3–6 pellagra epidemic exposure |  |  | -0.036***<br>(-0.050 to -0.022) |  | -0.032<br>(-0.066 to 0.002) |
| Age 7–10 pellagra epidemic exposure |  |  |  | -0.038***<br>(-0.054 to -0.022) | -0.006<br>(-0.036 to 0.025) |
| Marginal R<sup>2</sup> / Conditional R<sup>2</sup> | 0.31 / 0.64 | 0.31 / 0.64 | 0.31 / 0.64 | 0.31 / 0.64 | 0.31 / 0.64 |
| AIC | 487029 | 487030 | 487023 | 487026 | 487044 |
| N people<sup>a</sup> | 13708 | 13708 | 13708 | 13708 | 13708 |
| N observations | 94414 | 94414 | 94414 | 94414 | 94414 |

\* $p \leq 0.05$ \*\* $p \leq 0.01$ \*\*\* $p \leq 0.001$
Abbreviations: AD: Alzheimer's disease; AIC: Akaike information criterion; APOE: apolipoprotein E; N: number; PGS: polygenic score.
Notes: All models included a random intercept at the person level. Values presented are coefficient estimates with 95% confidence intervals. Models were adjusted for gender, age (cubic polynomial centered at 65), mother's education, participant's education, practice effects, childhood family financial situation, person's birth Census division, birth year cohort in 5-year intervals, and the first 10 principal components. Models 1–4 regress the age-based exposures of interest separately, whereas model 5 includes all of those exposures. *In utero* pellagra epidemic exposure refers to the pellagra mortality rate during a participant's gestation year in their state of birth. The age period pellagra exposures refer to the average of the pellagra mortality rates in the state of birth for each participant over the years a participant was a given age.
<sup>a</sup> For consistency, the sample size in these analyses was restricted to people who had exposure values across all age categories, which is fewer individuals than in the main analysis.

#### 1.3.5 Additional sensitivity analyses

Additional sensitivity analyses with other genetic measures, the global cognition 35-point measure, within the South Census region, using alternative specifications for location and birth period, without adjusting for participant’s education, and adjusting for health factors remained consistent in direction and close in magnitude with the main results (**Supplemental Results**).

### 1.4 DISCUSSION

Using a natural experiment based on the 20^th^ century pellagra epidemic, we found that early-life exposure to living through the pellagra epidemic was associated with late-life global cognition. The effect of *in utero* exposure to the epidemic was modest; model estimates predicted an approximate one-point difference in cognitive function on a 27-point scale between someone with no exposure and someone with high exposure. For reference, this difference is about the same as that between males and females in this sample. These findings are consistent with literature on the harmful effect of famines in early life on late-life cognition (de Rooij et al., 2010). However, famines are relatively large shocks compared to the relatively small shock of experiencing a more severe pellagra epidemic. The small effects of *in utero* pellagra epidemic exposure on late-life cognition are consistent with other studies of *in utero* effects on late-life outcomes (Furuya and Fletcher, 2022). We found that this observed effect on late-life cognition was not isolated to prenatal exposure to the epidemic. Exposure during age periods through age 10 were individually associated with late-life global cognition, but when evaluated together, no single age period stood out as an influence on cognition. Exposure during the prenatal and early-childhood period are correlated with each other because pellagra death rates gradually decreased over time, so it is difficult to separate the effects from one another. However, if the effect of one period is strong enough under these conditions, finding an effect is possible (Schmitz and Duque, 2022).

When stratified by race and gender, the direction and approximate magnitude of the exposure effect were consistent in most strata, even though the association was no longer significant in all but Black females and females of other racial backgrounds. The effect of the exposure in White male and female strata was the same as in the full sample, which is unsurprising because they compose most of the sample. The exposure effect was greatest in Black females and females from other racial background groups. Historically disadvantaged groups were most vulnerable to the pellagra epidemic and had higher rates of death from pellagra, so lower cognition levels in late-life would be consistent with a more severe experience of the epidemic (Marks, 2003). Although not significant, the effect of the exposure was positive in Black men and men from other racial background groups, which may reflect accelerated aging and high mortality selection experienced by these groups (Levine and Crimmins, 2014). This mortality selection increases the likelihood that Black people and men from other racial groups who survived to be in the HRS and who remained in the study are healthier than those who did not.

However, neither of the effects for these two groups were significant, and the sample size for males of other racial backgrounds was less than 250. Overall, the stratified results do highlight both racial and gender differences in the effect of *in utero* pellagra epidemic exposure on late-life cognition that may reflect either selection or different conditions each group experienced within a particular exposure level.

We also evaluated whether the *in utero* exposure could modify the effect of AD genetic risk on cognitive function. We found a significant interaction between the AD PGS (with *APOE)* and *in utero* pellagra epidemic exposure that suggested that during a more severe pellagra epidemic there were greater differences in the effect of genetics on cognition. In other words, the negative environment amplified the effects of genetics or, equivalently, a positive environment lessened the effect of genetics. High risk genetic groups had worse cognition and low risk genetic groups had better cognition in worse *in utero* environments compared to better *in utero* environments. Sensitivity analyses revealed that the interaction effects were likely driven by *APOE* as the PGS without *APOE* did not have a significant interaction, but *APOE* alone did interact with *in utero* pellagra epidemic exposure. *APOE* driving the statistical association does not imply that any biological interaction can be solely attributed to *APOE*. Rather, the interaction analysis of the PGS without *APOE* may have been underpowered because of the PGSs weaker main effect. Overall, the results suggest that a better *in utero* environment relating to the pellagra epidemic can attenuate genetic effects on late-life cognition.

There were a few limitations with the main exposure in the study. First, the pellagra epidemic exposure was not a proxy for B-vitamin availability during the *in utero* period for HRS participants and was not a direct measure of the nutritional environment. Along the same line, everyone with a given *in utero* pellagra epidemic exposure did not experience the same level of nutritional deprivation. Second, we could only generate crude pellagra mortality rates as a measure of the pellagra epidemic and not ones adjusted for the underlying population for a state and year. For the same level of nutritional deprivation, one state may have had a higher pellagra mortality rate than another because its population had, for example, a higher percentage of females, who are at higher risk of dying from pellagra than males (Brenton, 2000). Third, accuracy of pellagra mortality statistics may have varied with pellagra epidemic severity. For example, there was a stigma associated with some areas that had high pellagra death rates and subsequent resistance to acknowledge the deaths occurring in the area (Bollet, 1992), which means the true effect of the *in utero* exposure could have been even stronger than reported. Despite the limitations, the approach of using a natural experiment was less prone to spurious associations and overcame measurement challenges like poor recall, which are normally associated with dietary studies. The natural experiment created by the pellagra epidemic was a rare opportunity to not only study the effects of an early-life factor on late-life cognition—an exposure and outcome separated by a considerable time span—but also to study a complex behavioral and social exposure like the nutritional environment.

Additional measurement aspects should be considered when interpreting the results of this study. The cognitive score used in the analysis is less able to distinguish cognitive differences in the middle range of cognition (e.g., cognitively impaired non-demented) than it is in the range of normal cognition or post-dementia diagnosis (Crimmins et al., 2011). However, it was available for a large number of individuals, allowing us to uncover the relatively small but important effect of an *in utero* exposure. In addition, the global cognition score was more heavily weighted on memory cognitive domains, which means that the effect of *in utero* pellagra mortality rate on cognition was largely an effect on memory aspects of cognition. Fully understanding the effects of the *in utero* exposure on late-life cognition would benefit from an evaluation of multiple cognitive domains. The PGSs used in the analysis were a few of several available for measuring genetic risk of AD and have varying degrees of accuracy in predicting a particular outcome (Escott-Price et al., 2015; Escott-Price and Hardy, 2022).

Although the HRS is a nationally representative sample, the analytic sample is not because many individuals were missing key measures and were removed. In addition to generalizability considerations, readers should keep in mind that survival bias may affect the results. With an *in utero* exposure, selection into live birth is a potential issue for inference of the effects of that prenatal exposure (Bruckner and Catalano, 2018; Nobles and Hamoudi, 2019). If the harmful effects of high *in utero* pellagra epidemic exposure selected against more frail individuals who also would have had lower cognitive values in later life, the *in utero* effects presented in our study would be biased downward.

There is also potential survival bias regarding entry into HRS. To be eligible for the study, individuals had to have survived to retirement age. If a high *in utero* pellagra epidemic exposure also affected survival, there would be fewer of those individuals surviving to study entry. This would result in another bias that attenuated the effects of the *in utero* exposure on cognition. The subsample of HRS participants with genetic data is even more susceptible to survival bias because only those who survived to 2006, when genetic sampling was first conducted, were included.

The natural experiment created by the pellagra epidemic provided a unique opportunity to overcome measurement challenges associated with studying early-life nutritional effects on a late-life outcome such as cognition. Although the *in utero* effect was modest and not the sole early-life period associated with cognition, it did persist into late-life. The presence of an effect that carries over from early to late life gives credence to further expanding the window of time for studying changes relating to dementia. In addition, it highlights that nutritional environments are an important target for evaluating mechanisms in dementia progression.

## ACKNOWLEDGEMENTS

This work is supported by the National Institute on Aging (NIA) grants R01AG054047 and RF1AG054047. Eva Vasiljevic was supported by an NIA grant, T32 AG00129, awarded through the Center for Demography of Health and Aging. Computational resources for this research were supported by a core grant to the Center for Demography and Ecology at the University of Wisconsin-Madison (P2C HD047873). The HRS is sponsored by the NIA grant U01AG009740 and is conducted by the University of Michigan. RAND HRS data are supported with funding from the NIA and the Social Security Administration. The authors thank Katherine Brenneke for her aid in mortality file organization and data entry and Daniel Panyard and Jaime Preussler for reviewing the manuscript for readability.

## 1.5 DATA AVAILABILITY

Health and Retirement Study (HRS) restricted and sensitive individual-level data were used in this study. We are not allowed to share those data because of our agreement with HRS and because research participant privacy could be compromised. Instructions for accessing HRS data can be found at https://hrs.isr.umich.edu/data-products. Pellagra death counts from the US Census Bureau are publicly available through the National Center for Health Statistics at https://www.cdc.gov/nchs/products/vsus/vsus_1890_1938.htm and at https://www.cdc.gov/nchs/products/vsus/vsus_1939_1964.htm. Intercensal population estimates for each state are publicly available from the US Census Bureau https://www.census.gov/data/tables/time-series/demo/popest/pre-1980-state.html.

## 1.6 CODE AVAILABILITY

New software was not developed for this study. The software used for the study is available from the cited sources.

## SUPPLEMENTAL METHODS

### Sample and data

#### HRS description

The target population of the HRS is household-dwelling adults, aged 51–61, who live within the contiguous US. Household financial units with at least one member who was born between 1931 and 1941 were eligible for the original HRS sample. These observational units were selected with a national multi-stage area probability sample design. Black, Hispanic, and Floridian households were oversampled (Juster and Suzman, 1995). Within the observational unit, both the age-eligible adult and their spouse (if they had one) were interviewed to get individual-level and household-level characteristics (Fisher and Ryan, 2017). Additional cohorts were added to the original HRS sample, and now, the HRS uses a steady-state design to replenish the sample every six years with additional retirement cohorts (HRS Staff, 2008; Sonnega et al., 2014). The HRS assesses participants every two years (Fisher and Ryan, 2017). Interview mode includes face-to-face at baseline, telephone for interviews between 1994 and 2004, and alternating face-to-face with telephone modes thereafter (Fisher and Ryan, 2017). The alternating modes are carried out by interviewing half of the sample face-to-face and the other half over the telephone. The mode for an individual is flipped the following wave such that each person undergoes a face-to-face interview every four years.

#### HRS data sources

Public use datasets included the “Cross-Wave Tracker File” (April 2023, v3.0, Early), “Cross-Wave Childhood Health and Family Aggregated Data” (April 2020 v. 2.0), and “Polygenic Score Data” (February 2021, v4.3). Researcher-contributed public use datasets included the “Langa-Weir Classification of Cognitive Function” (May 2023) and the “RAND HRS Longitudinal File 2020” (May 2024, v2.0) produced by the RAND Center for the Study of Aging. Sensitive data used in the analyses came from the dataset “*APOE* and Serotonin Transporter Alleles” (August 2021, v1.0, Early). Restricted data came from “Cross-Wave Geographic Information (Detail) [1992–2020]” (April 2023, v9.0). Cleaned and imputed Individual-level genetic data for generating the polygenic scores (PGS) for HRS participants were obtained through the database of genotypes and phenotypes repository (dbGaP, accession number: phs000428.v2.p2, phases 1–3, 2006–2010 samples).

### Measures

#### Exposure: Exposure to the pellagra epidemic

Pellagra mortality rates per 100K for each state and year were derived from two data sources: death counts for each year and US state and US Census Bureau population estimates for those states and years. Intercensal population estimates for each state are publicly available and were obtained from the US Census Bureau (US Census Bureau, 2018). Pellagra death counts assembled by the US Census Bureau in the early 20th century are publicly available and were obtained through the National Center for Health Statistics (Centers For Disease Control, National Center for Health Statistics, 1939, 1890–1938). Mortality tables were provided in portable document format (pdf) files from scanned mortality reports. Pellagra death counts from 1915 through and inclusive of 1950 were manually entered for all states that reported pellagra deaths. For a full list of tables see **Supplemental Table 1**. To check for data entry errors, pellagra death counts were plotted over time for each state and outliers were examined against original reports for correctness. In addition, the sum of state death counts was calculated and compared to the US total of pellagra deaths provided in the mortality statistics. If the state and US total did not match, the entered state death counts for pellagra were double checked and corrected. If no data entry errors were found, the state and US pellagra death counts were subsequently compared to another tabulation of pellagra deaths in the mortality reports if it was available. Mismatching state and US totals were resolved using these two methods in all but one year, 1934. For 1934, the state total (3600) was off by 2 from the US total (3602). It was assumed there was an error in the original report for this year. To calculate the mortality rate, death counts for a state and year were divided by that state’s estimated population for that year and multiplied by 100K (see mortality rates across years for each state in **Supplemental Figure 1**).

In addition, three periods of exposure to the pellagra epidemic during childhood were included. These childhood age stages were 0 through 2 years, 3 through 6 years, and 7 through 10 years. In particular, the 0–2 year period approximates the “first 1000 days of life” and is a sensitive period of child development that tends to be the focus of research and the target of health policies (Suri et al., 2025). There are no clear standards for dividing childhood into different age groups as development periods vary depending on the specific developmental factor of interest. Therefore, we divided the remaining years of childhood into two equal periods. The age stage exposures were calculated by averaging state-and year-level pellagra mortality rates through their first, second, and third years of life for the 0 through 2 year period, for example. For example, for a child born in April of 1932 in South Carolina, their first year of life exposure would be equivalent to the weighted average of the pellagra mortality rates in South Carolina for 1932 and 1933 based on the fraction of months lived in each year assuming an end of month birthdate, 8/12 and 4/12 for this case, respectively. The measures assume children stayed in their state of birth through childhood.

#### Outcome: Global cognition

The 27-point global cognition is a scale that sums the following tests: immediate and delayed 10-noun free recall test to measure memory (immediate and delayed memory, scale: 0–20); a sevens subtraction test (working memory, scale: 0–5); and a counting backwards test (processing speed, scale: 0–2).

Imputation of individual tests and more details on the 27-point cognition are already described (Langa et al., 2022; McCammon et al., 2019).

For sensitivity analyses, the 35-point global cognition measure was used. It includes the following additional questions that were only administered to participants over age 65: date naming (scale: 0–4); object naming (scale: 0–2); and naming the US president and the vice president (scale: 0–2). This measure was available in the RAND longitudinal file.

#### Genetic measures

The PGS for an individual was calculated as the weighted mean across the included variants, where the number of risk alleles for a variant were weighted by the effect size of that variant’s association with AD (Wray et al., 2007). Weights in this score primarily came from the Kunkle et al. (2019) GWAS for AD, which was conducted in individuals of European genetic ancestry. Variants used for the PGS came from individual-level genetic data from dbGaP except for the *APOE* variants, which were obtained from the higher-quality genotyping data provided for those variants in HRS’s sensitive survey data (using the TaqMan assay instead of the genotyping array) (Faul et al., 2021). Weights for *APOE* variants came from the Kunkle et al. AD GWAS (2019). Among individuals with imputed *APOE* values, we took the additional step of removing individuals with *APOE* variant imputation quality scores below 0.8.

Quality control procedures for the dbGaP-obtained individual-level data have already been described (Health and Retirement Study, 2013a) as have the data imputation procedures (Health and Retirement Study, 2013b). Imputed individual-level data were provided as allele probabilities, which were converted in PLINK to hard calls using a 90% posterior probability threshold.

The de Rojas et al. (2021) summary statistics for the PGS were harmonized with HRS genetic data by conforming all variants to the same DNA reference strand. The effect estimate in the summary statistics was reversed (multiplied by −1) if the HRS genetic data were coded using the opposite reference strand (e.g., AG and GA). Ambiguous allele combinations (AT, TA, GC, and CG) were resolved by matching alleles based on the reference allele frequencies in the summary statistics and in allele frequencies in HRS.

Weights for the HRS-provided AD PGSs came from the 2013 International Genomics of Alzheimer’s Project GWAS (Lambert et al., 2013). The HRS AD PGS was obtained from publicly available HRS data and slightly modified as described below. Additional details about all HRS PGS construction are included in HRS documentation (Ware et al., 2019). HRS provides two AD PGSs, one with and one without *APOE* variants (rs7412 and rs429358), and these PGSs are generated using genotyping array data. We substituted the *APOE* variants in the HRS-provided AD PGS with the ones from the higher quality measurement technique by taking the HRS AD PGS that does not include *APOE* and adding the weighted better quality *APOE* variants (as described for the main AD PGS). Unlike the main analysis AD PGSs, the HRS-provided AD PGSs were calculated as a weighted sum and not a weighted mean of the variants in the PGS. The advantage of the weighted mean used in the main analysis over a sum is that individuals missing values for genetic variants will not necessarily have lower PGS values.

The HRS-provided cognition PGS was based on weights from a GWAS conducted in data from the Cohorts for Heart and Aging Research in Genomic Epidemiology consortium (Davies et al., 2015). Higher values of the PGS for cognition indicate better cognition. Additional details for this PGS are provided in HRS documentation (Ware et al., 2019).

#### Covariates

Categorical covariates were coded using the following categories: gender (female = 1, male = 0), race (Black or African American, White or Caucasian [reference], other racial backgrounds [aggregate category provided by HRS]), participant’s education (less than high school [HS], general education diploma [GED], HS [reference], some college, college and above), mother’s education (no schooling [reference], less than 8 years, completed 8 years, high school graduate, above high school, missing) childhood family financial situation (poor, average [reference], well off, varied, missing), birth year cohort (five year interval categories starting in 1915 and going through 1954 [reference: 1915–1919]), and birth Census division (East North Central [reference], West North Central, New England, Middle Atlantic, West South Central, East South Central, South Atlantic, Pacific, Mountain), and interview language (English [reference] or Spanish). Practice effect was calculated at each cognitive assessment as the number of previous cognitive assessments available for that person. All categorical covariates were coded as nominal variables (i.e., not ordinal) with the indicated reference category. A sensitivity analysis adjusted for the following indicators of ever having or experiencing (never = 0, ever = 1) a stroke, hypertension, heart disease or problems, diabetes, and smoking. These indicators came from the RAND longitudinal dataset.

### Statistical analysis

#### Regression modeling

In addition to a random slope at the individual level to account for repeated measures correlation, we tested if a random slope for age at the individual level would improve fit over the random-intercept-only model using the Akaike Information Criterion and an F-test with the “anova” function in R (R Core Team, 2020). Although it did significantly improve the fit, the random slope for age caused convergence issues even after trying different optimization functions. Therefore, we did not include the random slope for age in any models.

Regression diagnostics were performed for all models, which included evaluating the mean structure and heteroscedasticity using residual-versus-fitted-value plots and evaluating normality using quantile-quantile plots. Model estimates were estimated using restricted maximum likelihood. All models used an unstructured variance-covariance structure. The Satterthwaite approximation was used to determine degrees of freedom, and 0.05 was the alpha significance criterion.

## SUPPLEMENTAL RESULTS

### Genetic sensitivity analyses

The pattern observed with the main analysis AD PGS was also observed using HRS-provided PGSs for AD with and without *APOE*. The interaction was significant when *APOE* was included in the AD (β = −0.010, 95% CI: −0.020 to 0.000, **Supplemental Table 5**) and not significant when it was excluded (β = −0.005, 95% CI: −0.017 to 0.008, **Supplemental Table 5**). The interaction with the cognition PGS was not significant (β = −0.003, 95% CI: −0.015 to 0.008, **Supplemental Table 5**). **Global cognition 35-point outcome**

Associations of *in utero* pellagra epidemic exposure with the 35-point global cognition outcome were consistent with those using the 27-point cognitive outcome. Although the effect sizes were not directly comparable between the two cognitive outcomes, they were similar in magnitude. In the full sample not restricted to participants with genetic data, the effect of *in utero* pellagra epidemic exposure was significant (β = −0.033, 95% CI: −0.044 to −0.022, model 1, **Supplemental Table 6**). The interaction between the AD PGS with *APOE* and the *in utero* pellagra epidemic exposure was also significant as it was in the models with the 27-point cognition (β = −2.457, 95% CI: −3.908 to −1.007, model 2b, **Supplemental Table 6**).

### Analysis within South Census region

The effect of the *in utero* pellagra epidemic exposure on 27-point cognition within a subset of individuals born in the US South Census region remained significant and was consistent in direction and magnitude with the effect in the full sample (β = −0.021, 95% CI: −0.033 to −0.008, **Supplemental Table 7**). The interaction between *in utero* pellagra epidemic exposure and the AD PGS was also consistent in direction and magnitude with the full analytic sample analysis. However, it was no longer significant (β = −1.714, 95% CI: −3.432 to 0.004, **Supplemental Table 7**).

### Alternative specifications for location and birth period

Associations between *in utero* pellagra epidemic exposure and 27-point global cognition remained significant in analyses including state of birth and birth year as fixed effects and in analyses including those terms as random intercepts (**Supplemental Table 8**). The effect of the exposure on global cognition in the analysis with fixed effects was not as strong as the effect in the analysis with random intercepts (β = −0.018 vs β = −0.024). The effect of *in utero* pellagra epidemic exposure in the main analysis (β = −0.025, **Table 2**) was essentially the same as in the model with a random intercept specification. The confidence intervals were almost identical as well.

### Analysis without participant’s education

*In utero* pellagra epidemic exposure remained significant and was consistent in direction with the effect in the full sample. However, the effect was slightly stronger when modeled without participant’s education than in the original model adjusting for education (β = −0.032, 95% CI: −0.043 to −0.021, **Supplemental Table 9**).

### Analysis including health factors

The association between *in utero* exposure to the pellagra epidemic remained unchanged after adjusting for history of stroke, hypertension, heart disease or problems, diabetes, and smoking (β = −0.025, 95% CI: −0.035 to −0.015, **Supplemental Table 10**). Likewise, the interaction between the AD PGS and *in utero* pellagra epidemic exposure remained significant after inclusion of these covariates (β = −1.921, 95% CI: −3.229 to −0.614) and essentially the same as the original model (original β = −1.915, **Table 3**). As previously noted, to generate the analytic sample, visits when stroke was first reported and all subsequent visits for that individual were removed. However, participants themselves were not removed based on stroke history.

## SUPPLEMENTAL FIGURES AND TABLES

**Supplemental Figure 1:**
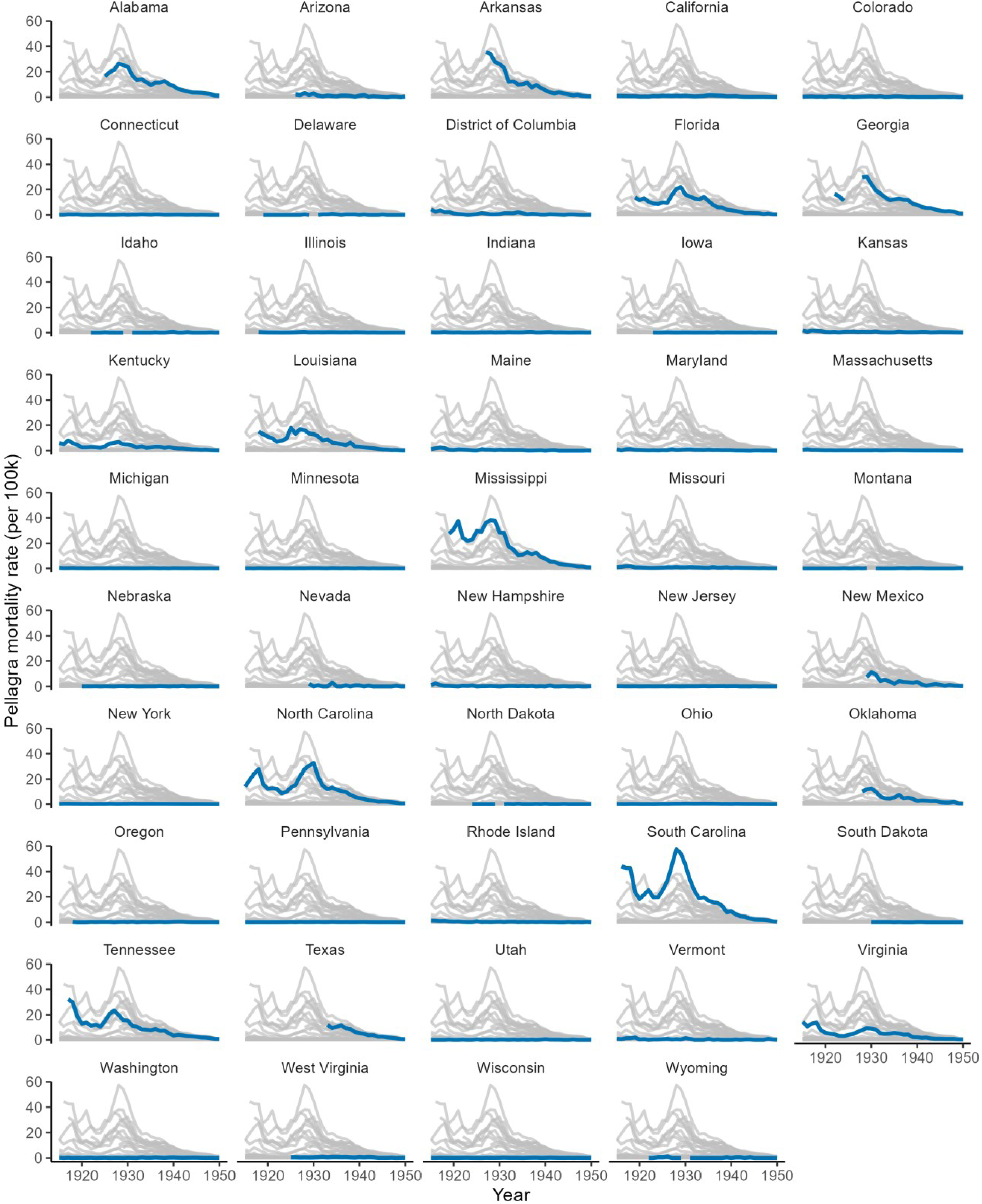
Pellagra mortality rates (y-axis) from 1915 through 1950 (x-axis) for US states and the District of Columbia. Each state’s pellagra mortality rate is plotted in a thick blue line with grey lines representing the mortality rates of all other states. Breaks in the line represent lack of data availability for a given year.

**Supplemental Figure 2:**
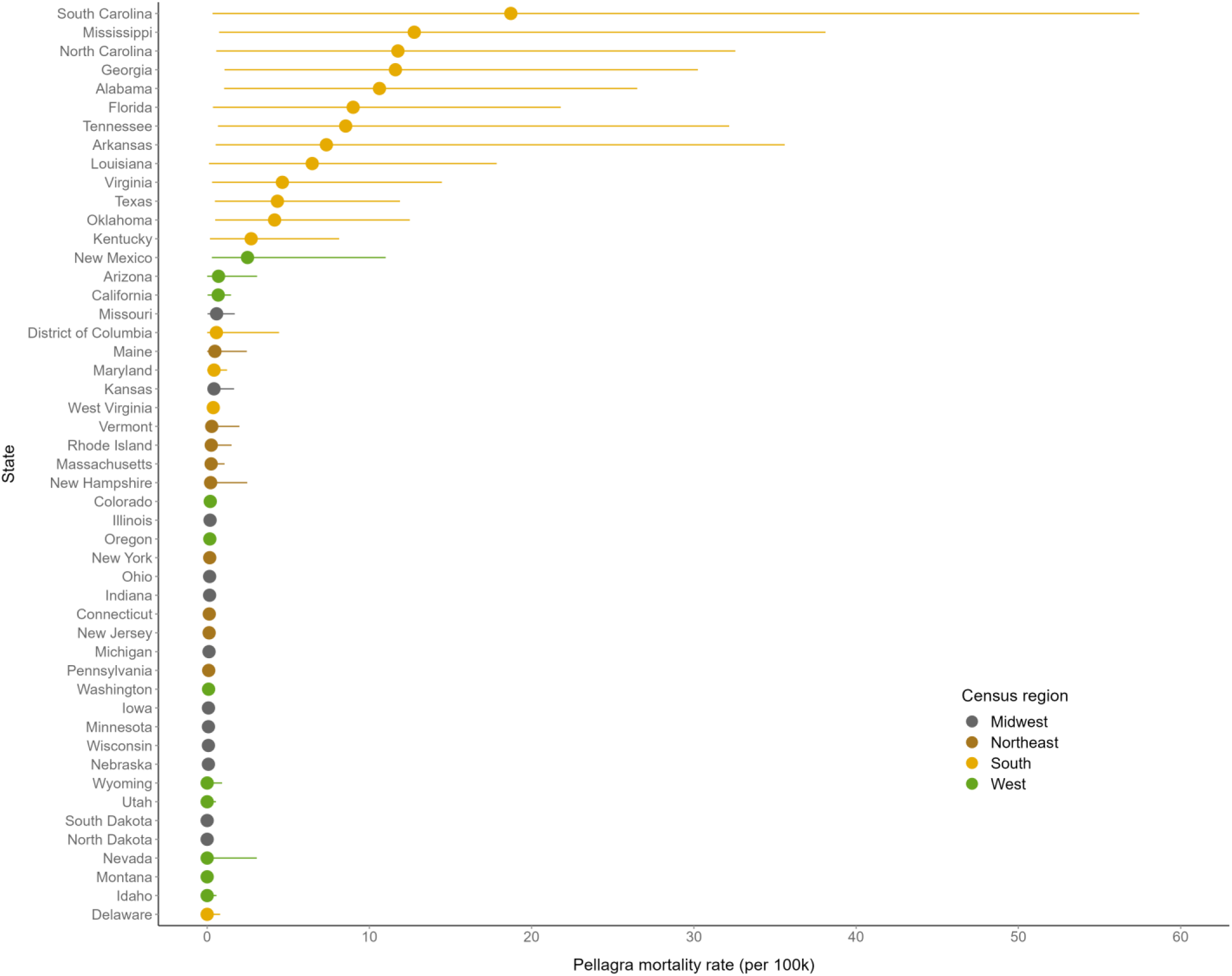
Range of pellagra mortality rates per 100,000 people in each state in the data from 1915 through 1950. The median pellagra mortality rate for each state is indicated with a point, and lines extending from the point indicate minimum and maximum values across available pellagra mortality data for that state. States are sorted by median mortality rate in descending order. Colors indicate the Census region a state belongs to.

**Supplemental Table 1:** List of mortality tables from which pellagra deaths were obtained.

| Year | Report title | VSUS filename | Table number in VSUS | Table title |
| --- | --- | --- | --- | --- |
| 1915 | Mortality Statistics 1915 | mortstatsh_1915.pdf | Table 6 | Deaths (exclusive of stillbirths) in the registration area, its main subdivisions, and in cities and rural districts of each registration state, from each cause and class of causes: 1915 |
| 1916 | Mortality Statistics 1916 | mortstatsh_1916.pdf | Table 6 | Deaths (exclusive of stillbirths) in the registration area, its main subdivisions, and in cities and rural districts of each registration state, from each cause and class of causes: 1916 |
| 1917 | Mortality Statistics 1917 | mortstatsh_1917.pdf | Table 6 | Deaths (exclusive of stillbirths) in the registration area (exclusive of Hawaii), its main subdivisions, and in cities and rural districts of each registration state, from each cause and class of causes: 1917 |
| 1918 | Mortality Statistics 1918 | mortstatsh_1918.pdf | Table 6 | Deaths (exclusive of stillbirths) in the registration area (exclusive of Hawaii), its main subdivisions, and in cities and rural districts of each registration state, from each cause and class of causes: 1918 |
| 1919 | Mortality Statistics 1919 | mortstatsh_1919.pdf | Table 6 | Deaths (exclusive of stillbirths) in the registration area (exclusive of Hawaii), its main subdivisions, and in cities and rural districts of each registration state, from each cause and class of causes: 1919 |
| 1920 | Mortality Statistics 1920 | mortstatsh_1920.pdf | Table 6 | Deaths (exclusive of stillbirths) in the registration area (exclusive of Hawaii), its main subdivisions, and in cities and rural districts of each registration state, from each cause and class of causes: 1920 |
| 1921 | Mortality Statistics 1921 | mortstatsh_1921.pdf | Table 6 | Deaths (exclusive of stillbirths) in the registration area (exclusive of Hawaii), its main subdivisions, and in cities and rural districts of each registration state, from each cause and class of causes: 1921 |
| 1922 | Mortality Statistics 1922 | mortstatsh_1922.pdf | Table 6 | Deaths (exclusive of stillbirths) in the registration area (exclusive of Hawaii), its main subdivisions, and in cities and rural districts of each registration state, from each cause and class of causes: 1922 |
| 1923 | Mortality Statistics 1923 | mortstatsh_1923.pdf | Table 6 | Deaths (exclusive of stillbirths) from each cause, in urban and rural districts: 1923 |
| 1924 | Mortality Statistics 1924 | mortstatsh_1924.pdf | Table 6 | Deaths (exclusive of stillbirths) from each cause, in urban and rural districts: 1924 |
| 1925 | Mortality Statistics 1925, Part 1 | mortstatsh_1925.pdf | Table 6 | Deaths (exclusive of stillbirths) from each cause, urban and rural districts: 1925 |
| 1926 | Mortality Statistics 1926, Part 1 | mortstatsh_1926.pdf | Table 6 | Deaths (exclusive of stillbirths) from each cause, in urban and rural districts: 1926 |
| 1927 | Mortality Statistics 1927, Part 1 | mortstatsh_1927.pdf | Table 6 | Deaths (exclusive of stillbirths) from each cause, in urban and rural districts: 1927 |
| 1928 | Mortality Statistics 1928 | mortstatsh_1928.pdf | Table 6 | Deaths (exclusive of stillbirths) from each cause, in urban and rural districts: 1928 |
| 1929 | Mortality Statistics 1929 | mortstatsh_1929.pdf | Table 6 | Deaths (exclusive of stillbirths) from each cause, in urban and rural districts: 1929 |

| **Year**<br>**(Continued)** | **Report title** | **VSUS filename** | **Table**<br>**number in**<br>**VSUS** | **Table title** |
| --- | --- | --- | --- | --- |
| **1930** | Mortality Statistics 1931 | mortstatsh_1931.pdf | Table AN | No table name provided |
| **1931** | Mortality Statistics 1931 | mortstatsh_1931.pdf | Table 8 | Deaths from each cause in cities and rural areas and by age, for the registration area and each registration state: 1931 |
| **1932** | Mortality Statistics 1932 | mortstatsh_1932.pdf | Table 8 | Deaths (exclusive of stillbirths) from each cause in cities and rural areas and by age for the registration area and each registration state: 1932 |
| **1933** | Mortality Statistics 1933 | mortstatsh_1933.pdf | Table 8 | Deaths (exclusive of stillbirths) from each cause in cities and rural areas, and by age, in continental United States and each state: 1933 |
| **1934** | Mortality Statistics 1934 | mortstatsh_1934.pdf | Table 6 | Deaths (exclusive of stillbirths) from each cause, in each state, by cities and rural districts: 1934 |
| **1935** | Mortality Statistics 1935 | mortstatsh_1935.pdf | Table 6 | Deaths (exclusive of stillbirths) from each cause, in each state, by cities and rural districts: 1935 |
| **1936** | Mortality Statistics 1936 | mortstatsh_1936.pdf | Table 6 | Deaths (exclusive of stillbirths) from each cause, in each state, by cities and rural areas: 1936 |
| **1937** | Vital Statistics of the United States 1937, Part 1 | VSUS_1937_1.pdf | Table 17 | Deaths from each cause by race and sex: United States and each state, 1937 |
| **1938** | Vital Statistics of the United States 1938, Part 1 | VSUS_1938_1.tmp | Table 18 | Deaths from each cause by race and sex: United States and each state, 1938 |
| **1939** | Vital Statistics of the United States 1939, Part 1 | VSUS_1939_1.pdf | Table 14 | Deaths from each cause by race and sex: United States and each state, 1939 |
| **1940** | Vital Statistics of the United States 1940, Part 1 | vsus_1940_1.pdf | Table 15 | Deaths from each cause, by race and sex: United States and each State, 1940 |
| **1941** | Vital Statistics of the United States 1941, Part 1 | vsus_1941_1.pdf | Table 9 | Deaths from each cause, by race and sex: United States and each State, 1941 |
| **1942** | Vital Statistics of the United States 1942, Part 2 | vsus_1942_2.pdf | Table 24 | Deaths from each cause, by race and sex: United States and each State, 1942 |
| **1943** | Vital Statistics of the United States 1943, Part 2 | vsus_1943_2.pdf | Table 24 | Deaths from each cause, by race and sex: United States and each State, 1943 |
| **1944** | Vital Statistics of the United States 1944, Part 2 | vsus_1944_2.pdf | Table 24 | Deaths from each cause, by race and sex: United States and each State, 1944 |
| **1945** | Vital Statistics of the United States 1945, Part 2 | vsus_1945_2.pdf | Table 19 | Deaths from each cause, by race and sex: United States and each State, 1945 |
| **1946** | Vital Statistics of the United States 1946, Part 2 | vsus_1946_2.pdf | Table 21 | Deaths from each cause, by race and sex: United States and each State, 1946 |
| **1947** | Vital Statistics of the United States 1947, Part 2 | vsus_1947_2.pdf | Table 22 | Deaths from each cause, by race and sex: United States and each State, 1947 |
| **1948** | Vital Statistics of the United States 1948, Part 2 | vsus_1948_2.pdf | Table 22 | Deaths from each cause, by race and sex: United States and each State, 1948 |
| **1949** | Vital Statistics of the United States 1949, Part 2 | vsus_1949_2.pdf | Table 23 | Deaths from 254 selected causes, by race and sex: United States and each State, 1949 |
| **1950** | Vital Statistics of the United States 1950, Volume 3 | vsus_1950_3.pdf | Table 54 | Deaths from 255 selected causes, by race and sex: United States and each State, 1950 |

**Supplemental Table 2:**
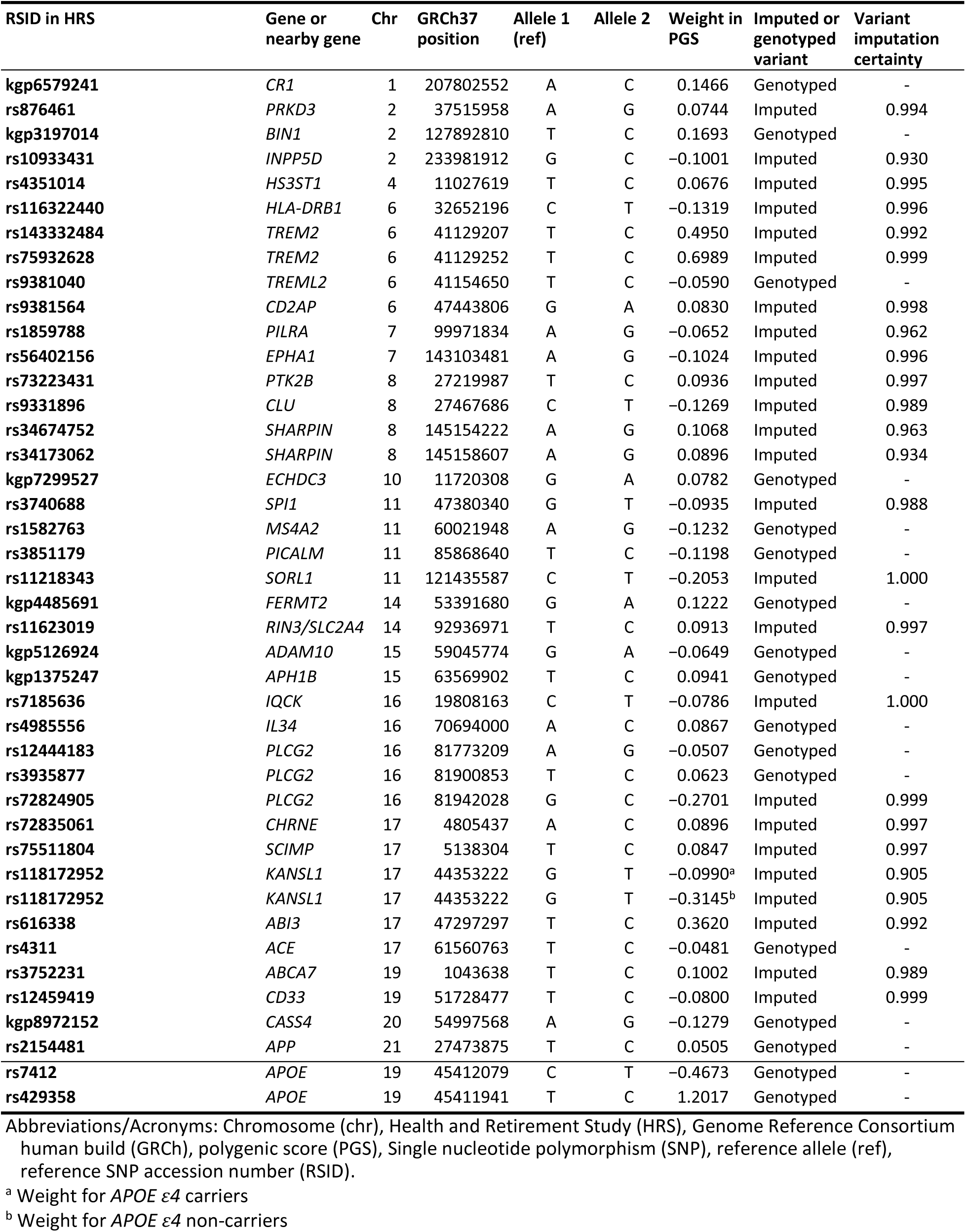
List of variants in the main analysis polygenic score.

| RSID in HRS | Gene or nearby gene | Chr | GRCh37 position | Allele 1 (ref) | Allele 2 | Weight in PGS | Imputed or genotyped variant | Variant imputation certainty |
| --- | --- | --- | --- | --- | --- | --- | --- | --- |
| kgp6579241 | CR1 | 1 | 207802552 | A | C | 0.1466 | Genotyped | - |
| rs876461 | PRKD3 | 2 | 37515958 | A | G | 0.0744 | Imputed | 0.994 |
| kgp3197014 | BIN1 | 2 | 127892810 | T | C | 0.1693 | Genotyped | - |
| rs10933431 | INPP5D | 2 | 233981912 | G | C | -0.1001 | Imputed | 0.930 |
| rs4351014 | HS3ST1 | 4 | 11027619 | T | C | 0.0676 | Imputed | 0.995 |
| rs116322440 | HLA-DRB1 | 6 | 32652196 | C | T | -0.1319 | Imputed | 0.996 |
| rs143332484 | TREM2 | 6 | 41129207 | T | C | 0.4950 | Imputed | 0.992 |
| rs75932628 | TREM2 | 6 | 41129252 | T | C | 0.6989 | Imputed | 0.999 |
| rs9381040 | TREML2 | 6 | 41154650 | T | C | -0.0590 | Genotyped | - |
| rs9381564 | CD2AP | 6 | 47443806 | G | A | 0.0830 | Imputed | 0.998 |
| rs1859788 | PILRA | 7 | 99971834 | A | G | -0.0652 | Imputed | 0.962 |
| rs56402156 | EPHA1 | 7 | 143103481 | A | G | -0.1024 | Imputed | 0.996 |
| rs73223431 | PTK2B | 8 | 27219987 | T | C | 0.0936 | Imputed | 0.997 |
| rs9331896 | CLU | 8 | 27467686 | C | T | -0.1269 | Imputed | 0.989 |
| rs34674752 | SHARPIN | 8 | 145154222 | A | G | 0.1068 | Imputed | 0.963 |
| rs34173062 | SHARPIN | 8 | 145158607 | A | G | 0.0896 | Imputed | 0.934 |
| kgp7299527 | ECHDC3 | 10 | 11720308 | G | A | 0.0782 | Genotyped | - |
| rs3740688 | SPI1 | 11 | 47380340 | G | T | -0.0935 | Imputed | 0.988 |
| rs1582763 | MS4A2 | 11 | 60021948 | A | G | -0.1232 | Genotyped | - |
| rs3851179 | PICALM | 11 | 85868640 | T | C | -0.1198 | Genotyped | - |
| rs11218343 | SORL1 | 11 | 121435587 | C | T | -0.2053 | Imputed | 1.000 |
| kgp4485691 | FERMT2 | 14 | 53391680 | G | A | 0.1222 | Genotyped | - |
| rs11623019 | RIN3/SLC2A4 | 14 | 92936971 | T | C | 0.0913 | Imputed | 0.997 |
| kgp5126924 | ADAM10 | 15 | 59045774 | G | A | -0.0649 | Genotyped | - |
| kgp1375247 | APH1B | 15 | 63569902 | T | C | 0.0941 | Genotyped | - |
| rs7185636 | IQCK | 16 | 19808163 | C | T | -0.0786 | Imputed | 1.000 |
| rs4985556 | IL34 | 16 | 70694000 | A | C | 0.0867 | Genotyped | - |
| rs12444183 | PLCG2 | 16 | 81773209 | A | G | -0.0507 | Genotyped | - |
| rs3935877 | PLCG2 | 16 | 81900853 | T | C | 0.0623 | Genotyped | - |
| rs72824905 | PLCG2 | 16 | 81942028 | G | C | -0.2701 | Imputed | 0.999 |
| rs72835061 | CHRNE | 17 | 4805437 | A | C | 0.0896 | Imputed | 0.997 |
| rs75511804 | SCIMP | 17 | 5138304 | T | C | 0.0847 | Imputed | 0.997 |
| rs118172952 | KANSL1 | 17 | 44353222 | G | T | -0.0990<sup>a</sup> | Imputed | 0.905 |
| rs118172952 | KANSL1 | 17 | 44353222 | G | T | -0.3145<sup>b</sup> | Imputed | 0.905 |
| rs616338 | ABI3 | 17 | 47297297 | T | C | 0.3620 | Imputed | 0.992 |
| rs4311 | ACE | 17 | 61560763 | T | C | -0.0481 | Genotyped | - |
| rs3752231 | ABCA7 | 19 | 1043638 | T | C | 0.1002 | Imputed | 0.989 |
| rs12459419 | CD33 | 19 | 51728477 | T | C | -0.0800 | Imputed | 0.999 |
| kgp8972152 | CASS4 | 20 | 54997568 | A | G | -0.1279 | Genotyped | - |
| rs2154481 | APP | 21 | 27473875 | T | C | 0.0505 | Genotyped | - |
| rs7412 | APOE | 19 | 45412079 | C | T | -0.4673 | Genotyped | - |
| rs429358 | APOE | 19 | 45411941 | T | C | 1.2017 | Genotyped | - |

Abbreviations/Acronyms: Chromosome (chr), Health and Retirement Study (HRS), Genome Reference Consortium human build (GRCh), polygenic score (PGS), Single nucleotide polymorphism (SNP), reference allele (ref), reference SNP accession number (RSID).
<sup>a</sup> Weight for APOE ε4 carriers
<sup>b</sup> Weight for APOE ε4 non-carriers

**Supplemental Table 3:** Association between *in utero* pellagra epidemic exposure and 27-point global cognition using mixed effects regression models listing all covariate estimates.

| Covariate | Outcome: 27-point global cognition |  |  |  |
| --- | --- | --- | --- | --- |
|  | Model 1 | Model 2 | Model 3a<sup>a</sup> | Model 3b<sup>a</sup> |
| Intercept | 15.618<br>(15.560 to 15.676) | 13.006<br>(12.761 to 13.251) | 11.992<br>(11.546 to 12.439) | 11.676<br>(11.239 to 12.112) |
| *In utero* pellagra epidemic exposure | -0.148***<br>(-0.156 to -0.139) | -0.059***<br>(-0.070 to -0.047) | -0.024***<br>(-0.033 to -0.014) | -0.025***<br>(-0.035 to -0.015) |
| Age (years, c = 65) |  |  | -0.213***<br>(-0.224 to -0.202) | -747.951***<br>(-784.477 to -711.425) |
| Age (2nd deg.) |  |  |  | -169.267***<br>(-175.691 to -162.844) |
| Age (3rd deg.) |  |  |  | -12.954***<br>(-18.951 to -6.956) |
| Gender (female) |  |  | 0.939***<br>(0.853 to 1.025) | 0.963***<br>(0.877 to 1.050) |
| Race: black (ref: white) |  |  | -2.030***<br>(-2.162 to -1.899) | -2.057***<br>(-2.189 to -1.924) |
| Race: other (ref: white) |  |  | -1.387***<br>(-1.666 to -1.108) | -1.410***<br>(-1.690 to -1.129) |
| Mother's edu: < 8yrs (ref: no school) |  |  | 1.392***<br>(1.030 to 1.754) | 1.380***<br>(1.016 to 1.744) |
| Mother's edu: 8yrs (ref: no school) |  |  | 1.770***<br>(1.411 to 2.129) | 1.757***<br>(1.396 to 2.118) |
| Mother's edu: HS (ref: no school) |  |  | 1.829***<br>(1.465 to 2.194) | 1.814***<br>(1.447 to 2.180) |
| Mother's edu: > HS (ref: no school) |  |  | 2.095***<br>(1.714 to 2.477) | 2.080***<br>(1.696 to 2.464) |
| Mother's edu: missing (ref: no school) |  |  | 1.035***<br>(0.663 to 1.407) | 1.005***<br>(0.631 to 1.379) |
| Edu: GED (ref: < HS) |  |  | 1.850***<br>(1.634 to 2.066) | 1.859***<br>(1.641 to 2.076) |
| Edu: HS (ref: < HS) |  |  | 2.484***<br>(2.359 to 2.608) | 2.502***<br>(2.377 to 2.627) |
| Edu: some college (ref: < HS) |  |  | 3.343***<br>(3.204 to 3.481) | 3.367***<br>(3.227 to 3.506) |
| Edu: college and above (ref: < HS) |  |  | 4.533***<br>(4.385 to 4.681) | 4.554***<br>(4.405 to 4.703) |
| Practice effects |  |  | 0.095***<br>(0.071 to 0.118) | 0.114***<br>(0.091 to 0.138) |
| Interview language: Spa (ref: Eng) |  |  | -0.057<br>(-0.522 to 0.408) | 0.039<br>(-0.424 to 0.502) |
| Childhood fam. fin.: well off (ref: average) |  |  | -0.165<br>(-0.348 to 0.018) | -0.162<br>(-0.346 to 0.022) |
| Childhood fam. fin.: poor (ref: average) |  |  | 0.135**<br>(0.038 to 0.231) | 0.145**<br>(0.048 to 0.241) |
| Childhood fam. fin.: varied (ref: average) |  |  | 0.220<br>(-0.199 to 0.639) | 0.190<br>(-0.232 to 0.612) |
| Childhood fam. fin.: missing (ref: average) |  |  | -0.162<br>(-0.445 to 0.121) | -0.188<br>(-0.471 to 0.095) |

| (Continued) |  | Outcome: 27-point global cognition |  |  |
| --- | --- | --- | --- | --- |
| Covariate | Model 1 | Model 2 | Model 3a<sup>a</sup> | Model 3b<sup>a</sup> |
| Birth division: WNC (ref: ENC) |  | 0.043<br>(-0.144 to 0.229) | -0.001<br>(-0.158 to 0.156) | -0.003<br>(-0.160 to 0.155) |
| Birth division: NE (ref: ENC) |  | -0.028<br>(-0.269 to 0.212) | -0.056<br>(-0.259 to 0.146) | -0.060<br>(-0.264 to 0.144) |
| Birth division: MA (ref: ENC) |  | 0.186*<br>(0.024 to 0.349) | 0.253***<br>(0.116 to 0.390) | 0.252***<br>(0.114 to 0.390) |
| Birth division: WSC (ref: ENC) |  | -1.267***<br>(-1.485 to -1.049) | -0.273**<br>(-0.461 to -0.085) | -0.270**<br>(-0.459 to -0.081) |
| Birth division: ESC (ref: ENC) |  | -1.230***<br>(-1.451 to -1.008) | -0.282**<br>(-0.471 to -0.092) | -0.281**<br>(-0.472 to -0.090) |
| Birth division: SA (ref: ENC) |  | -1.285***<br>(-1.467 to -1.104) | -0.279***<br>(-0.436 to -0.121) | -0.275***<br>(-0.433 to -0.116) |
| Birth division: P (ref: ENC) |  | -0.104<br>(-0.332 to 0.124) | -0.262**<br>(-0.454 to -0.069) | -0.264**<br>(-0.458 to -0.071) |
| Birth division: M (ref: ENC) |  | -0.775***<br>(-1.062 to -0.487) | -0.559***<br>(-0.804 to -0.315) | -0.570***<br>(-0.816 to -0.324) |
| Birth year cohort 1920–1924 (ref: 1915–1919) |  | 0.845***<br>(0.566 to 1.125) | 0.000<br>(-0.242 to 0.242) | -0.352**<br>(-0.595 to -0.109) |
| Birth year cohort 1925–1929 (ref: 1915–1919) |  | 1.623***<br>(1.354 to 1.893) | -0.112<br>(-0.359 to 0.136) | -0.664***<br>(-0.913 to -0.414) |
| Birth year cohort 1930–1934 (ref: 1915–1919) |  | 2.704***<br>(2.447 to 2.962) | -0.113<br>(-0.385 to 0.158) | -0.859***<br>(-1.133 to -0.585) |
| Birth year cohort 1935–1939 (ref: 1915–1919) |  | 3.204***<br>(2.950 to 3.457) | -0.508**<br>(-0.811 to -0.206) | -1.263***<br>(-1.568 to -0.958) |
| Birth year cohort 1940–1944 (ref: 1915–1919) |  | 3.776***<br>(3.518 to 4.035) | -0.930***<br>(-1.268 to -0.592) | -1.593***<br>(-1.933 to -1.253) |
| Birth year cohort 1945–1949 (ref: 1915–1919) |  | 4.028***<br>(3.763 to 4.294) | -1.387***<br>(-1.747 to -1.026) | -1.964***<br>(-2.326 to -1.602) |
| Birth year cohort 1950–1954 (ref: 1915–1919) |  | 3.529***<br>(3.199 to 3.858) | -1.751***<br>(-2.145 to -1.357) | -2.334***<br>(-2.730 to -1.939) |
| Marginal R<sup>2</sup> / Conditional R<sup>2</sup> | 0.03 / 0.56 | 0.09 / 0.56 | 0.30 / 0.62 | 0.32 / 0.64 |
| AIC | 687648 | 685895 | 664279 | 661621 |
| N people | 18285 | 18285 | 18285 | 18285 |
| N observations | 128927 | 128927 | 128927 | 128927 |

\* p ≤ 0.05 \*\* p ≤ 0.01 \*\*\* p ≤ 0.001
Abbreviations: AIC: Akaike information criterion; c: centered; deg: degree; N: number; ref: reference; HS: high school; GED: general education diploma; Spa: Spanish; Eng: English; ENC: East North Central; WNC: West North Central; NE: New England; MA: Middle Atlantic; WSC: West South Central; ESC: East South Central; SA: South Atlantic; P: Pacific; M: Mountain.
Notes: Values presented are coefficient estimates with 95% confidence intervals. All models included a random intercept at the person level. *In utero* pellagra epidemic exposure refers to the pellagra mortality rate during a participant's gestation year in their state of birth.
<sup>a</sup> Age is specified as a linear term in model 3a and as a cubic polynomial in model 3b.

**Supplemental Table 4:** Association between *in utero* pellagra epidemic exposure and 27-point cognition stratified by race and gender.

| Covariate | Outcome: 27-point global cognition |  |  |  |  |  |
| --- | --- | --- | --- | --- | --- | --- |
|  | White male | White female | Black male | Black female | Other male | Other female |
| **Intercept** | 11.923<br>(11.180 to 12.665) | 12.680<br>(11.964 to 13.395) | 9.460<br>(7.454 to 11.466) | 9.396<br>(7.805 to 10.987) | 8.471<br>(3.939 to 13.002) | 14.444<br>(10.578 to 18.310) |
| ***In utero* pellagra epidemic exposure** | -0.018<br>(-0.036 to 0.001) | -0.015<br>(-0.033 to 0.002) | 0.004<br>(-0.027 to 0.035) | -0.031*<br>(-0.059 to -0.004) | 0.061<br>(-0.072 to 0.194) | -0.145**<br>(-0.242 to -0.049) |
| **Age (years, c = 65)** | -424.886***<br>(-456.806 to -392.966) | -566.665***<br>(-610.739 to -522.590) | -133.320***<br>(-159.520 to -107.120) | -199.166***<br>(-235.468 to -162.863) | -59.000***<br>(-85.832 to -32.167) | -79.065***<br>(-112.353 to -45.777) |
| **Age (2nd deg.)** | -86.218***<br>(-92.309 to -80.126) | -136.350***<br>(-142.810 to -129.890) | -23.095***<br>(-29.589 to -16.601) | -39.272***<br>(-46.390 to -32.154) | -13.657***<br>(-20.010 to -7.304) | -20.264***<br>(-27.133 to -13.396) |
| **Age (3rd deg.)** | -1.822<br>(-7.533 to 3.889) | -9.024**<br>(-15.045 to -3.003) | -6.794*<br>(-12.899 to -0.688) | -4.325<br>(-11.006 to 2.356) | 0.342<br>(-5.610 to 6.294) | -4.006<br>(-10.492 to 2.480) |
| **Mother's edu: < 8yrs (ref: no school)** | 1.132***<br>(0.477 to 1.786) | 1.514***<br>(0.874 to 2.153) | 1.483*<br>(0.178 to 2.788) | 1.869***<br>(0.895 to 2.843) | 0.421<br>(-1.329 to 2.172) | 0.299<br>(-1.185 to 1.784) |
| **Mother's edu: 8yrs (ref: no school)** | 1.416***<br>(0.773 to 2.059) | 2.057***<br>(1.424 to 2.690) | 1.279<br>(-0.023 to 2.582) | 2.094***<br>(1.119 to 3.070) | 1.378<br>(-0.474 to 3.231) | 0.603<br>(-1.020 to 2.226) |
| **Mother's edu: HS (ref: no school)** | 1.452***<br>(0.804 to 2.099) | 2.178***<br>(1.537 to 2.820) | 1.185<br>(-0.163 to 2.533) | 2.221***<br>(1.189 to 3.253) | 1.386<br>(-0.654 to 3.426) | 0.788<br>(-1.023 to 2.598) |
| **Mother's edu: > HS (ref: no school)** | 1.989***<br>(1.318 to 2.660) | 2.248***<br>(1.588 to 2.908) | 1.004<br>(-0.516 to 2.523) | 2.350***<br>(1.182 to 3.518) | 2.150<br>(-0.486 to 4.785) | suppressed<sup>a</sup> |
| **Mother's edu: missing (ref: no school)** | 0.877**<br>(0.214 to 1.541) | 0.971**<br>(0.312 to 1.630) | 0.815<br>(-0.510 to 2.140) | 1.726***<br>(0.719 to 2.733) | 0.807<br>(-1.081 to 2.696) | 0.059<br>(-1.651 to 1.769) |
| **Edu: GED (ref: < HS)** | 1.785***<br>(1.464 to 2.106) | 1.543***<br>(1.183 to 1.902) | 1.994***<br>(1.078 to 2.910) | 2.511***<br>(1.755 to 3.268) | 2.790**<br>(1.033 to 4.547) | 3.090***<br>(1.271 to 4.908) |
| **Edu: HS (ref: < HS)** | 2.433***<br>(2.222 to 2.645) | 2.317***<br>(2.131 to 2.503) | 2.732***<br>(2.231 to 3.233) | 2.823***<br>(2.419 to 3.226) | 2.472***<br>(1.133 to 3.812) | 2.327***<br>(1.050 to 3.604) |
| **Edu: some college (ref: < HS)** | 3.164***<br>(2.935 to 3.392) | 3.134***<br>(2.923 to 3.346) | 3.896***<br>(3.338 to 4.453) | 4.140***<br>(3.670 to 4.609) | 4.366***<br>(2.997 to 5.736) | 3.818***<br>(2.534 to 5.102) |
| **Edu: college and above (ref: < HS)** | 4.462***<br>(4.236 to 4.689) | 4.169***<br>(3.936 to 4.403) | 5.741***<br>(5.011 to 6.471) | 5.634***<br>(5.091 to 6.178) | 5.100***<br>(3.477 to 6.722) | 6.095***<br>(4.365 to 7.825) |
| **Practice effects** | 0.128***<br>(0.093 to 0.163) | 0.121***<br>(0.082 to 0.159) | 0.099*<br>(0.013 to 0.185) | 0.077<br>(-0.004 to 0.159) | 0.187<br>(-0.007 to 0.380) | 0.137<br>(-0.055 to 0.329) |
| **Childhood fam. fin.: well off (ref: average)** | -0.210<br>(-0.493 to 0.073) | -0.173<br>(-0.435 to 0.088) | 0.287<br>(-0.766 to 1.340) | -0.111<br>(-0.964 to 0.742) | 0.091<br>(-1.720 to 1.901) | 0.711<br>(-1.168 to 2.591) |

| (Continued) |  |  |  |  |  |  |
| --- | --- | --- | --- | --- | --- | --- |
| Covariate | Outcome: 27-point global cognition |  |  |  |  |  |
|  | White male | White female | Black male | Black female | Other male | Other female |
| Childhood fam. fin.: poor (ref: average) | 0.248**<br>(0.095 to 0.400) | 0.083<br>(-0.064 to 0.231) | 0.205<br>(-0.191 to 0.602) | -0.024<br>(-0.353 to 0.305) | -0.644<br>(-1.646 to 0.358) | 0.287<br>(-0.647 to 1.221) |
| Childhood fam. fin.: varied (ref: average) | 0.497<br>(-0.139 to 1.132) | 0.091<br>(-0.499 to 0.680) | suppressed<sup>a</sup> | suppressed<sup>a</sup> |  | suppressed<sup>a</sup> |
| Childhood fam. fin.: missing (ref: average) | -0.355<br>(-0.775 to 0.066) | -0.026<br>(-0.474 to 0.422) | -0.153<br>(-1.252 to 0.946) | -0.087<br>(-1.090 to 0.915) | suppressed<sup>a</sup> | suppressed<sup>a</sup> |
| Birth division: WNC (ref: ENC) | -0.044<br>(-0.271 to 0.183) | 0.008<br>(-0.211 to 0.226) | 1.362<br>(-0.296 to 3.020) | -0.053<br>(-1.277 to 1.171) | suppressed<sup>a</sup> | suppressed<sup>a</sup> |
| Birth division: NE (ref: ENC) | 0.097<br>(-0.193 to 0.386) | -0.190<br>(-0.472 to 0.092) | suppressed<sup>a</sup> | suppressed<sup>a</sup> | suppressed<sup>a</sup> | suppressed<sup>a</sup> |
| Birth division: MA (ref: ENC) | 0.331**<br>(0.128 to 0.534) | 0.152<br>(-0.041 to 0.344) | 0.694<br>(-0.310 to 1.697) | 0.495<br>(-0.304 to 1.294) | 2.232<br>(-0.125 to 4.590) | suppressed<sup>a</sup> |
| Birth division: WSC (ref: ENC) | -0.249<br>(-0.568 to 0.071) | -0.412**<br>(-0.701 to -0.122) | -0.139<br>(-1.090 to 0.812) | -0.197<br>(-0.962 to 0.567) | 1.222<br>(-0.773 to 3.216) | -1.001<br>(-2.889 to 0.887) |
| Birth division: ESC (ref: ENC) | -0.334*<br>(-0.661 to -0.006) | -0.319*<br>(-0.619 to -0.018) | -0.512<br>(-1.448 to 0.423) | -0.062<br>(-0.795 to 0.670) | suppressed<sup>a</sup> | -2.051<br>(-4.617 to 0.515) |
| Birth division: SA (ref: ENC) | -0.254<br>(-0.513 to 0.005) | -0.281*<br>(-0.521 to -0.041) | -0.600<br>(-1.476 to 0.276) | -0.017<br>(-0.700 to 0.666) | -0.118<br>(-2.246 to 2.011) | -0.791<br>(-2.796 to 1.214) |
| Birth division: P (ref: ENC) | -0.160<br>(-0.450 to 0.130) | -0.345*<br>(-0.614 to -0.076) | -0.298<br>(-2.124 to 1.529) | 0.319<br>(-1.111 to 1.748) | 0.267<br>(-1.624 to 2.157) | -1.295<br>(-3.191 to 0.602) |
| Birth division: M (ref: ENC) | -0.340<br>(-0.714 to 0.034) | -0.939***<br>(-1.294 to -0.584) | suppressed<sup>a</sup> | suppressed<sup>a</sup> | 1.206<br>(-0.621 to 3.033) | -0.736<br>(-2.655 to 1.183) |
| Birth year cohort 1920–1924 (ref: 1915–1919) | -0.275<br>(-0.655 to 0.105) | -0.504**<br>(-0.839 to -0.168) | -0.004<br>(-1.455 to 1.446) | 0.006<br>(-1.181 to 1.192) | suppressed<sup>a</sup> | suppressed<sup>a</sup> |
| Birth year cohort 1925–1929 (ref: 1915–1919) | -0.768***<br>(-1.158 to -0.378) | -0.703***<br>(-1.054 to -0.351) | -0.255<br>(-1.694 to 1.185) | -0.005<br>(-1.205 to 1.194) | 1.233<br>(-2.854 to 5.319) | -2.044<br>(-5.206 to 1.119) |
| Birth year cohort 1930–1934 (ref: 1915–1919) | -1.016***<br>(-1.444 to -0.589) | -0.827***<br>(-1.232 to -0.421) | -0.589<br>(-1.996 to 0.818) | -0.526<br>(-1.714 to 0.662) | 1.209<br>(-2.999 to 5.417) | -1.604<br>(-4.874 to 1.666) |
| Birth year cohort 1935–1939 (ref: 1915–1919) | -1.348***<br>(-1.819 to -0.877) | -1.443***<br>(-1.910 to -0.975) | -0.924<br>(-2.397 to 0.548) | -0.197<br>(-1.478 to 1.084) | 0.502<br>(-3.686 to 4.690) | -3.098<br>(-6.534 to 0.338) |
| Birth year cohort 1940–1944 (ref: 1915–1919) | -1.660***<br>(-2.177 to -1.143) | -1.821***<br>(-2.356 to -1.287) | -0.845<br>(-2.423 to 0.733) | -0.761<br>(-2.155 to 0.633) | 1.089<br>(-3.333 to 5.511) | -3.357<br>(-7.068 to 0.354) |

| (Continued) |  | Outcome: 27-point global cognition |  |  |  |  |
| --- | --- | --- | --- | --- | --- | --- |
| Covariate | White male | White female | Black male | Black female | Other male | Other female |
| **Birth year cohort 1945–1949**<br>(ref: 1915–1919) | –2.030***<br>(–2.581 to –1.480) | –2.071***<br>(–2.653 to –1.489) | –1.473<br>(–3.079 to 0.133) | –1.262<br>(–2.698 to 0.174) | –0.155<br>(–4.621 to 4.311) | –4.112*<br>(–7.879 to –0.345) |
| **Birth year cohort 1950–1954**<br>(ref: 1915–1919) | –2.416***<br>(–3.040 to –1.793) | –2.505***<br>(–3.142 to –1.869) | –1.628<br>(–3.324 to 0.068) | –1.673*<br>(–3.156 to –0.190) | –0.814<br>(–5.504 to 3.877) | –3.964<br>(–7.950 to 0.023) |
| **Marginal R<sup>2</sup> / Conditional R<sup>2</sup>** | 0.28 / 0.61 | 0.27 / 0.61 | 0.29 / 0.65 | 0.30 / 0.63 | 0.32 / 0.68 | 0.33 / 0.65 |
| **AIC** | 224870 | 322555 | 33712 | 64031 | 6548 | 9291 |
| **N people** | 6731 | 8289 | 1108 | 1699 | 212 | 246 |
| **N observations** | 44715 | 62681 | 6513 | 11982 | 1275 | 1761 |

\* $p \leq 0.05$ \*\* $p \leq 0.01$ \*\*\* $p \leq 0.001$
Abbreviations: AIC: Akaike information criterion; c: centered; deg: degree; N: number; ref: reference; HS: high school; GED: general education diploma; ENC: East North Central; WNC: West North Central; NE: New England; MA: Middle Atlantic; WSC: West South Central; ESC: East South Central; SA: South Atlantic; P: Pacific; M: Mountain.
Notes: All models included a random intercept at the person level. Values presented are coefficient estimates with 95% confidence intervals. *In utero* pellagra epidemic exposure refers to the pellagra mortality rate during a participant's gestation year in their state of birth.
<sup>a</sup> Value is suppressed to mask an estimate based on fewer than 11 individuals.

**Supplemental Table 5:** Effect estimates of interaction between *in utero* pellagra epidemic exposure and alternative genetic measures.

| Genetic measure used in model | G x E interaction in model | Intercept | *In utero* exposure | Genetic measure | Genetic measure x *in utero* | Marginal R<sup>2</sup>/Conditional R<sup>2</sup> | AIC | N people | N observations |
| --- | --- | --- | --- | --- | --- | --- | --- | --- | --- |
| **AD PGS without *APOE*** | No | 13.762***<br>(12.723 to 14.801) | -0.010<br>(-0.027 to 0.006) | -10.957<br>(-23.366 to 1.452) |  | 0.27 / 0.58 | 357649 | 7625 | 71250 |
|  | Yes | 13.768***<br>(12.729 to 14.807) | -0.013<br>(-0.030 to 0.005) | -8.530<br>(-21.931 to 4.871) | -1.288<br>(-3.973 to 1.397) | 0.27 / 0.58 | 357647 | 7625 | 71250 |
| ***APOE*** | No | 13.872***<br>(12.838 to 14.906) | -0.011<br>(-0.028 to 0.005) | -0.381***<br>(-0.469 to -0.292) |  | 0.27 / 0.58 | 357590 | 7625 | 71250 |
|  | Yes | 13.857***<br>(12.823 to 14.891) | -0.004<br>(-0.021 to 0.013) | -0.325***<br>(-0.421 to -0.228) | -0.028**<br>(-0.047 to -0.009) | 0.27 / 0.58 | 357592 | 7625 | 71250 |
| **AD PGS with *APOE* interacted with PCs<sup>a</sup>** | Additional interaction with PCs | 13.783***<br>(12.749 to 14.817) | -0.014<br>(-0.042 to 0.014) | -21.708***<br>(-28.393 to -15.022) | -1.909**<br>(-3.235 to -0.583) | 0.27 / 0.58 | 357572 | 7625 | 71250 |
| **HRS-provided AD PGS with *APOE*** | No | 13.652***<br>(12.664 to 14.640) | -0.005<br>(-0.021 to 0.011) | -0.171***<br>(-0.216 to -0.125) |  | 0.27 / 0.58 | 379888 | 8136 | 75683 |
|  | Yes | 13.666***<br>(12.678 to 14.654) | -0.002<br>(-0.018 to 0.014) | -0.150***<br>(-0.199 to -0.101) | -0.010*<br>(-0.020 to 0.000) | 0.27 / 0.58 | 379895 | 8136 | 75683 |
| **HRS-provided AD PGS without *APOE*** | No | 13.601***<br>(12.610 to 14.591) | -0.005<br>(-0.021 to 0.011) | -0.100***<br>(-0.156 to -0.045) |  | 0.27 / 0.58 | 379930 | 8136 | 75683 |
|  | Yes | 13.605***<br>(12.615 to 14.596) | -0.005<br>(-0.021 to 0.011) | -0.091**<br>(-0.152 to -0.031) | -0.005<br>(-0.017 to 0.008) | 0.27 / 0.58 | 379939 | 8136 | 75683 |
| **HRS-provided cognition PGS** | No | 13.666***<br>(12.700 to 14.632) | -0.003<br>(-0.018 to 0.013) | 0.636***<br>(0.575 to 0.696) |  | 0.29 / 0.58 | 379527 | 8136 | 75683 |
|  | Yes | 13.668***<br>(12.702 to 14.635) | -0.003<br>(-0.018 to 0.013) | 0.642***<br>(0.578 to 0.707) | -0.003<br>(-0.015 to 0.008) | 0.29 / 0.58 | 379538 | 8136 | 75683 |

\* $p \leq 0.05$ \*\* $p \leq 0.01$ \*\*\* $p \leq 0.001$
Abbreviations: AD: Alzheimer's disease; AIC: Akaike information criterion; *APOE*: apolipoprotein E; N: number; PGS: polygenic score.
Notes: Each row represents a model of global cognition (the 27-point measure) regressed on the *in utero* pellagra epidemic exposure and a genetic measure.
*In utero* pellagra epidemic exposure refers to the pellagra mortality rate during a participant's gestation year in their state of birth. The column "Genetic measure used in model" indicates the genetic measure used in the regression instead of the AD PGS with *APOE* that was used in the main analysis, and the effect estimate of that measure is reported in the "genetic measure" column. The "G x E interaction in model" column indicates whether the model included an interaction between the indicated genetic measure and the *in utero* pellagra epidemic exposure, of which the effect estimate is reported in the "genetic measure x *in utero*" column. All models included a random intercept at the person level. Values presented are coefficient estimates with 95% confidence intervals. Models were adjusted for gender, age (cubic polynomial centered at 65), mother's education, participant's education, practice effects, childhood family financial situation, person's birth Census division, birth year cohort in 5-year intervals, and the first 10 principal components.
<sup>a</sup> In addition to the interaction between the AD PGS (with *APOE*) and the *in utero* pellagra epidemic exposure, the AD PGS is interacted with each of the 10 PCs in the model.

**Supplemental Table 6:** Association between *in utero* pellagra epidemic exposure and 35-point global cognition.

| Covariate | Outcome: 35-point global cognition |  |  |
| --- | --- | --- | --- |
|  | Model 1<sup>a</sup> | Model 2a<sup>b</sup> | Model 2b<sup>c</sup> |
| Intercept | 19.095<br>(18.578 to 19.612) | 21.801<br>(20.632 to 22.971) | 21.793<br>(20.624 to 22.962) |
| *In utero* pellagra epidemic exposure | -0.033***<br>(-0.044 to -0.022) | -0.014<br>(-0.032 to 0.005) | -0.010<br>(-0.029 to 0.009) |
| AD PGS with *APOE* |  | -34.537***<br>(-41.609 to -27.465) | -29.294***<br>(-37.009 to -21.578) |
| PGS x *in utero* |  |  | -2.457***<br>(-3.908 to -1.007) |
| Marginal R<sup>2</sup> / Conditional R<sup>2</sup> | 0.34 / 0.66 | 0.28 / 0.60 | 0.28 / 0.60 |
| AIC | 457457 | 244044 | 244034 |
| N people | 17949 | 7556 | 7556 |
| N observations | 85436 | 46823 | 46823 |

\* $p \leq 0.05$ \*\* $p \leq 0.01$ \*\*\* $p \leq 0.001$
Abbreviations: AD: Alzheimer's disease; AIC: Akaike information criterion; *APOE*: apolipoprotein E; N: number; PGS: polygenic score.
Notes: All models included a random intercept at the person level. Values presented are coefficient estimates with 95% confidence intervals. *In utero* pellagra epidemic exposure refers to the pellagra mortality rate during a participant's gestation year in their state of birth.
<sup>b</sup> Model 1 is the association of *in utero* pellagra epidemic exposure with 35-point global cognition in the full sample. It is adjusted for gender, race, age (cubic polynomial centered at 65), participant's education, mother's education, practice effects, interview language, childhood family financial situation, person's birth Census division, and birth year cohort in 5-year intervals.
<sup>c</sup> Models 2a and 2b show the association of the AD PGS on global cognition and the interaction of the PGS with *in utero* pellagra epidemic exposure. Covariates in these models are the same as in model 1 except that they do not adjust for race and interview language and do adjust for the first 10 principal components.

**Supplemental Table 7:** Association of *in utero* pellagra epidemic exposure with 27-point global cognition in a stratum of individuals born in the South Census region.

| Covariate | Outcome: 27-point global cognition<br>South Census region stratum |  |  |
| --- | --- | --- | --- |
|  | Model 1<sup>a</sup> | Model 2a<sup>b</sup> | Model 2b<sup>b</sup> |
| Intercept | 10.910<br>(10.154 to 11.665) | 13.824<br>(11.639 to 16.008) | 13.752<br>(11.568 to 15.936) |
| *In utero* pellagra epidemic exposure | -0.021**<br>(-0.033 to -0.008) | -0.025*<br>(-0.046 to -0.004) | -0.022*<br>(-0.043 to 0.000) |
| AD PGS with *APOE* |  | -35.169***<br>(-47.065 to -23.272) | -24.060**<br>(-40.349 to -7.770) |
| PGS x *in utero* |  |  | -1.714<br>(-3.432 to 0.004) |
| Marginal R<sup>2</sup> / Conditional R<sup>2</sup> | 0.35 / 0.66 | 0.27 / 0.59 | 0.27 / 0.59 |
| AIC | 238463 | 95701 | 95697 |
| N people | 6721 | 2079 | 2079 |
| N observations | 45876 | 18922 | 18922 |

\* $p \leq 0.05$ \*\* $p \leq 0.01$ \*\*\* $p \leq 0.001$
Abbreviations: AD: Alzheimer's disease; AIC: Akaike information criterion; *APOE*: apolipoprotein E; N: number; PGS: polygenic score.
All models included a random intercept at the person level. Values presented are coefficient estimates with 95% confidence intervals. *In utero* pellagra epidemic exposure refers to the pellagra mortality rate during a participant's gestation year in their state of birth.
<sup>a</sup> Model 1 is the association of *in utero* pellagra epidemic exposure with 27-point cognition in the full sample. It is adjusted for gender, race, age (cubic polynomial centered at 65), participant's education, mother's education, practice effects, interview language, childhood family financial situation, person's birth Census division, and birth year cohort in 5-year intervals.
<sup>b</sup> Models 2a and 2b show the association of the AD PGS on global cognition and the interaction of the PGS with *in utero* pellagra epidemic exposure. Covariates in these models are the same as in model 1 except that they do not adjust for race and interview language and do adjust for the first 10 principal components.

**Supplemental Table 8:** Association of *in utero* pellagra epidemic exposure with 27-point global cognition using alternative specifications of location and birth period.

| Outcome: 27-point global cognition |  |  |  |  |
| --- | --- | --- | --- | --- |
| Covariate | State of birth and birth year<br>included as fixed effects |  | State of birth and birth year<br>included as random intercepts |  |
|  | Base<sup>a</sup> | Fully-adjusted<sup>b</sup> | Base<sup>a</sup> | Fully-adjusted<sup>b</sup> |
| Intercept | 16.419<br>(16.043 to 16.794) | 10.606<br>(10.111 to 11.102) | 14.834<br>(14.266 to 15.402) | 10.541<br>(10.118 to 10.963) |
| *In utero*<br>pellagra<br>epidemic<br>exposure | -0.015*<br>(-0.029 to -0.002) | -0.018**<br>(-0.030 to -0.006) | -0.026***<br>(-0.040 to -0.013) | -0.024***<br>(-0.034 to -0.014) |
| Marginal R<sup>2</sup> /<br>Conditional R<sup>2</sup> | 0.10 / 0.56 | 0.32 / 0.64 | 0.00 / 0.58 | 0.34 / 0.66 |
| AIC | 685718 | 661735 | 685822 | 661685 |
| N people | 18285 | 18285 | 18285 | 18285 |
| N observations | 128927 | 128927 | 128927 | 128927 |

\* $p \leq 0.05$ \*\* $p \leq 0.01$ \*\*\* $p \leq 0.001$
Abbreviations: AIC: Akaike information criterion; N: number.
Notes: Values presented are coefficient estimates with 95% confidence intervals. All models included a random intercept at the person level. *In utero* pellagra epidemic exposure refers to the pellagra mortality rate during a participant's gestation year in their state of birth.
<sup>a</sup> Base models include state of birth and birth year as either covariates or random intercepts.
<sup>b</sup> Fully adjusted models are also adjusted for gender, race, age (cubic polynomial centered at 65), participant's education, mother's education, practice effects, and childhood family financial situation.

**Supplemental Table 9:** Association between *in utero* pellagra epidemic exposure and 27-point global cognition with and without adjusting for participant’s education.

|  | Outcome: 27-point global cognition |  |
| --- | --- | --- |
|  | With education | Without education |
| **Intercept** | 11.676<br>(11.239 to 12.112) | 13.223<br>(12.748 to 13.698) |
| ***In utero* pellagra epidemic exposure** | -0.025***<br>(-0.035 to -0.015) | -0.032***<br>(-0.043 to -0.021) |
| **Marginal R<sup>2</sup> / Conditional R<sup>2</sup>** | 0.32 / 0.64 | 0.25 / 0.64 |
| **AIC** | 661621 | 665081 |
| **N people** | 18285 | 18285 |
| **N observations** | 128927 | 128927 |

\* $p \leq 0.05$ \*\* $p \leq 0.01$ \*\*\* $p \leq 0.001$
Abbreviations: AIC: Akaike information criterion; c: centered; N: number.
Notes: Values presented are coefficient estimates with 95% confidence intervals. The models included a random intercept at the person level and adjusted for person's birth Census division, birth year cohort in 5-year intervals, gender, race, age (cubic polynomial centered at 65), mother's education, practice effects, interview language, and childhood family financial situation. *In utero* pellagra epidemic exposure refers to the pellagra mortality rate during a participant's gestation year in their state of birth.

**Supplemental Table 10:** Association of *in utero* pellagra epidemic exposure with 27-point cognition adjusting for health history.

| Covariates | Outcome: 27-point global cognition |  |  |
| --- | --- | --- | --- |
|  | Model 1<sup>a</sup> | Model 2a<sup>b</sup> | Model 2b<sup>b</sup> |
| Intercept | 11.904<br>(11.464 to 12.345) | 13.989<br>(12.953 to 15.024) | 13.986<br>(12.951 to 15.021) |
| *In utero* pellagra epidemic exposure | -0.025***<br>(-0.035 to -0.015) | -0.011<br>(-0.027 to 0.006) | -0.008<br>(-0.024 to 0.009) |
| AD PGS with APOE |  | -25.464***<br>(-31.587 to -19.340) | -21.590***<br>(-28.255 to -14.926) |
| PGS x *in utero* |  |  | -1.921**<br>(-3.229 to -0.614) |
| Stroke (ever) | -0.616**<br>(-1.001 to -0.231) | -0.339<br>(-0.826 to 0.148) | -0.343<br>(-0.830 to 0.143) |
| Hypertension (ever) | -0.157***<br>(-0.214 to -0.101) | -0.139***<br>(-0.210 to -0.067) | -0.139***<br>(-0.211 to -0.067) |
| Heart disease or problems (ever) | -0.133***<br>(-0.195 to -0.072) | -0.137***<br>(-0.215 to -0.059) | -0.137***<br>(-0.215 to -0.059) |
| Diabetes (ever) | -0.199***<br>(-0.267 to -0.131) | -0.185***<br>(-0.276 to -0.095) | -0.185***<br>(-0.276 to -0.094) |
| Smoke (ever) | -0.095*<br>(-0.184 to -0.007) | -0.079<br>(-0.196 to 0.038) | -0.081<br>(-0.198 to 0.036) |
| Marginal R<sup>2</sup> / Conditional R<sup>2</sup> | 0.32 / 0.64 | 0.27 / 0.58 | 0.27 / 0.58 |
| AIC | 657113 | 355263 | 355256 |
| N people | 18163 | 7580 | 7580 |
| N observations | 128092 | 70803 | 70803 |

\* $p \leq 0.05$ \*\* $p \leq 0.01$ \*\*\* $p \leq 0.001$
Abbreviations: AD: Alzheimer's disease; AIC: Akaike information criterion; APOE: apolipoprotein E; N: number; PGS: polygenic score.
Notes: All models included a random intercept at the person level. Values presented are coefficient estimates with 95% confidence intervals. *In utero* pellagra epidemic exposure refers to the pellagra mortality rate during a participant's gestation year in their state of birth.
<sup>a</sup> Model 1 is the association of *in utero* pellagra epidemic exposure with 27-point cognition in the full sample. It is adjusted for gender, race, age (cubic polynomial centered at 65), participant's education, mother's education, practice effects, interview language, childhood family financial situation, person's birth Census division, and birth year cohort in 5-year intervals.
<sup>b</sup> Models 2a and 2b show the association of the AD PGS on global cognition and the interaction of the PGS with *in utero* pellagra epidemic exposure. Covariates in these models are the same as in model 1 except that they do not adjust for race and interview language and do adjust for the first 10 principal components.

